# Causal effect of serum 25 hydroxyvitamin D concentration on cardioembolic stroke: Evidence from Two-sample Mendelian randomization

**DOI:** 10.1101/2023.10.11.23296798

**Authors:** Danial Habibi, Farshad Teymoori, Navid Ebrahimi, Sahand Tehrani Fateh, Leila Najd-Hassan-Bonab, Amir Hossein Saeidian, Alireza Soleymani Taloubaghi, Sara Asgarian, Farhad Hosseinpanah, Hakon Hakonarson, Fereidoun Azizi, Mehdi Hedayati, Maryam Sadat Daneshpour, Mahdi Akbarzadeh, Marjan Mansourian

## Abstract

**Background/Aim:** The putative association between serum 25 hydroxyvitamin D concentration 25(OH)D and the risk of cardioembolic stroke (CES) has been examined in observational studies, which indicate controversial findings. We performed Mendelian randomization (MR) analysis to determine the causal relationship of serum 25(OH)D with the risk of CES.

**Method:** The summary statistics dataset on the genetic variants related to 25(OH)D was used from the published GWAS of European descent participants in the UK Biobank, including 417,580 subjects, yielding 143 independent loci in 112 1-Mb regions. GWAS summary data of CES was obtained from GIGASTROKE Consortium, which included European individuals (10,804 cases, 1,234,808 controls).

**Results:** Our results unveiled 99 SNPs contributing a causal relationship between 25(OH)D and CES using IVW [OR□=□0.82, 95% CI: 0.67-0.98, p□=□0.037]. Horizontal pleiotropy was not seen by the MR-Egger intercept-based test [MR-Egger intercept□=□0.001; p□=□0.792], suggesting an absence of horizontal pleiotropy. Cochrane’s Q value [Q=78.71, p-value□=□0.924], Rucker’s Q [Q=78.64, p-value=0.913], and I^2^=0.0% (95% CI: 0.0%, 24.6%) statistic suggested no heterogeneity in the connection between 25(OH)D and CES. This result remained consistent using different MR method and sensitivity analyses, including Maximum likelihood [OR=0.82, 95%CI: 0.67-0.98, p-value=0.036], Constrained maximum likelihood method [OR=0.76, 95%CI: 0.64-0.90, p-value=0.002], Debiased inverse-variance weighted method [OR=0.82, 95%CI: 0.68-0.99, p-value=0.002], MR-PRESSO [OR=0.82, 95%CI 0.77-0.87, p-value=0.022], RAPS [OR=0.82, 95%CI 0.67-0.98, p-value=0.038], MR-Lasso [OR=0.82, 95%CI 0.68-0.99, p-value=0.037].

**Conclusion:** Our MR analysis provides suggestive evidence that increased 25(OH)D levels may play a causally protective role in the development of cardioembolic stroke. Determining the role of 25(OH)D in stroke subtypes has important clinical and public health implications.

## Introduction

Strokes are among the most common neurological disorders, major cause of death globally and considerable long-term disability(1). Ischemic stroke (IS) and hemorrhage stroke (HS) are both associated with high mortality(2). Prevalence of vascular risk factors and accordingly, the incidence of stroke, have likely influenced by lifestyle alterations and medical technology progressions during the past decade. Population aging and age-related risk factors, like atrial fibrillation (AF), may have influenced the incidence of cardioembolic stroke (CES)(3). Ischemic stroke can have different underlying causes, including conditions like atherosclerosis affecting cerebral blood vessels, occlusion of small cerebral vessels, and cardiac embolism. Accurate classification of ischemic stroke subtypes requires integration of clinical presentation, neuroimaging, cardiac, and vascular evaluation(4). Based on TOAST classification system, ischemic stroke comprises five categories including cardioembolic stroke. This category includes patients with arterial occlusions presumably due to an embolus with cardiac origin(5). Cardio-embolism is one of the most common etiologic causes of ischemic stroke based on several multicenter and single center demographic studies(6,7). While the overall incidence of stroke has decreased, there has been a threefold increase in the occurrence of cardioembolic strokes over the past few decades. Projections from the United Kingdom suggest that this trend could triple once more by the year 2050 (8).

A study in patients admitted with diagnosis of stroke showed that stroke patients had significantly lower serum vitamin D compared with healthy individuals(9). An observational study in China revealed that 25(OH)D and incidence of ischemic stroke are associated in an inverse-dose response manner(10). Additionally, it has been shown that vitamin D and its synthetic analogs, exerts anticoagulant effects via upregulation in expression of thrombomodulin (TM), an anticoagulant glycoprotein, and downregulation of tissue factor (TF), which could decrease thrombogenic activity of monocytes/macrophages(11). A systematic review and meta-analysis revealed that lower serum vitamin D measures is associated with an increased incidence of CES (12).

The link between vitamin D and ischemic stroke may be mediated by promotive effects of vitamin D on reendothelialization and angiogenesis. Vitamin D possess direct vascular effects and protecting vascular endothelial function (13).

Vitamin D deficiency is considered to be a global issue(14). The most common cause of vitamin D deficiency is reduced skin synthesis which can be related to aging and environmental conditions(15). In general population, the prevalence of vitamin D insufficiency is estimated to be 42% (16). Cardioembolic stroke (CES) is excessively more disabling than other mechanisms of stroke and increasing trend of ischemic stroke can be attributed to CES and might increase over the next decades in aging societies (17). Given the heavy social and economic burden, understanding the underlying mechanisms and evidence supporting causal role of vitamin D in CES would have important public health implications.

Mendelian randomization (MR) is a technique widely implemented to investigate the causality relationships between risk factors and various diseases in the absence of pleiotropy (18). This method uses genetic variants consistently associated with exposures of concern to estimate causal relationship between a given biomarker, such as vitamin D, and disease. This type of study implements assumptions that makes them less prone to reverse causation since disease states generally do not alter the germline DNA sequences. Moreover, due to random assortment of genotypes at meiosis, MR can limit confounding(18). Since genetic variants continue to be stable over a lifetime, MR method provide judicable evidence from a lifetime of genetically altered biomarker levels, for instance, lowered vitamin D levels. Consequently, MR methods can be compared to that of an RCT because the random assortment of genetic variants imitates the random allocation of participants to distinct therapeutic groups.

Since associations of genetically predicted 25-hydroxyvitamin D deficiency and cardioembolic stroke have not been unarguably established as previous studies have concerned analysis of total stroke, in the present study, we performed MR of genetically decreased serum 25-hydroxyvitamin D levels causal association with cardioembolic stroke.

## Research Design and Methods

### Study Design

This two-sample Mendelian Randomization study was designed to explore the causal effect of serum 25(OH)D concentration (exposure) on cardioembolic stroke (outcome) from European ancestry, as shown in Figure1. The study was based on publicly available summary-level data from genome-wide association studies (GWAS).

**Figure1:**
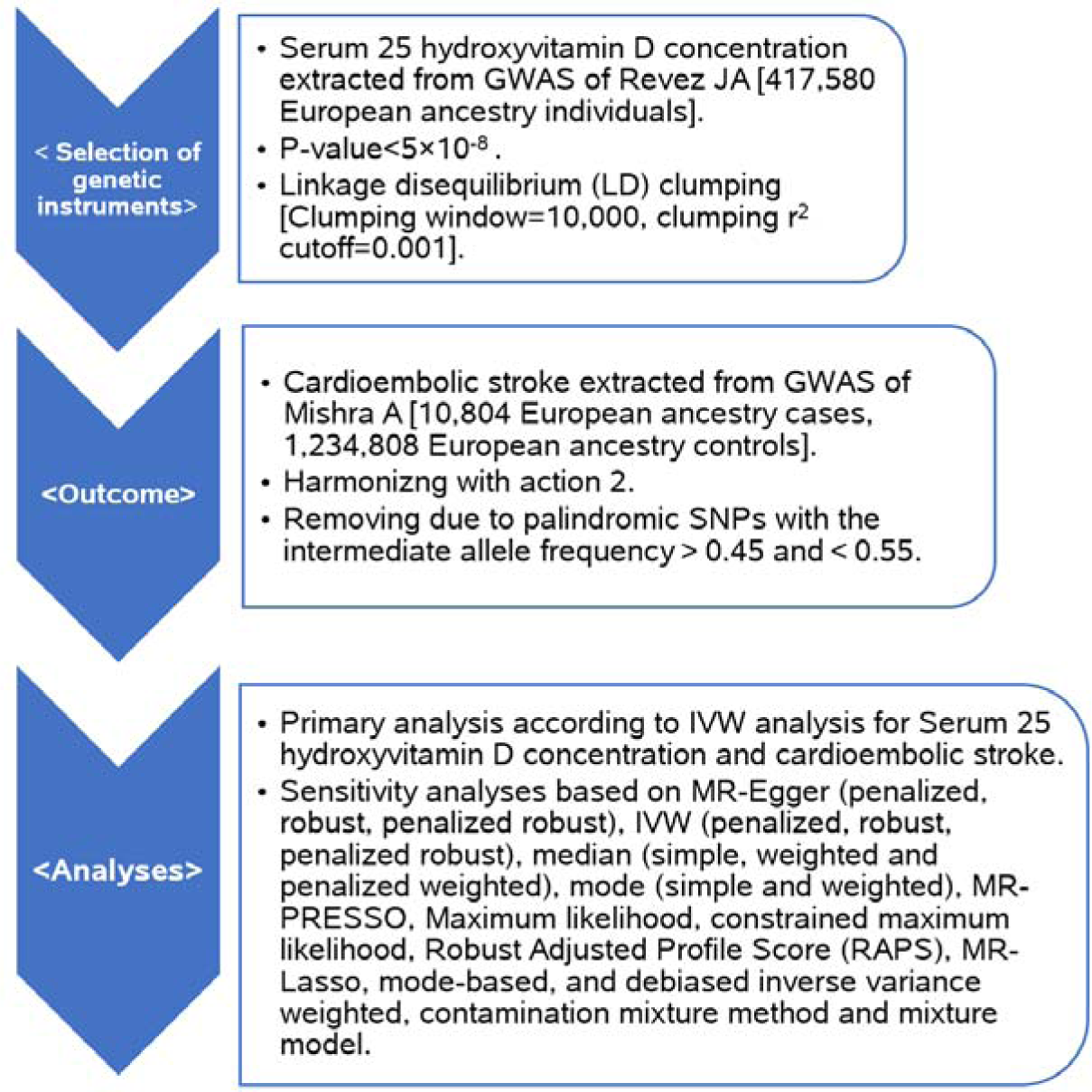
Mendelian Randomization design. Assessing the causal effect of serum 25 hydroxyvitamin D concentration on cardioembolic stroke.

### Data Sources

The dataset employed in this study was obtained from the GWAS catalog (https://www.ebi.ac.uk/gwas/). The current study utilized genetic associations derived from independent GWAS datasets with the same ancestral population to reduce the influence of confounding factors. Two sample Mendelian Randomization analysis was performed using published summary-level data in Table 1. Dataset of serum 25(OH)D concentration was a genome-wide association study in 417,580 Europeans with 143 independent loci in 112 1-Mb regions (19). For outcome, cardioembolic stroke dataset was including 10,804 European ancestry cases, and 1,234,808 European ancestry controls (20).

**Table 1:** Characteristics of Data Sources.

| Traits | PubMed ID | Author | Ancestor | Sample size | Use in this MR study |
| --- | --- | --- | --- | --- | --- |
| Vitamin D deficiency | 32,242,144 | Joana A. Revez | European | 417,580 | Exposure |
| cardioembolic stroke | 36,180,795 | Aniket Mishra | European | 10,804 cases,<br>1,234,808 controls | Outcome |

### Genetic Instrument Selection

To use Mendelian randomization, (i) Single nucleotide polymorphisms (SNPs) should be associated with the trait under study, the serum 25(OH)D concentration; (ii) SNPs be associated with the outcome through the serum 25(OH)D concentration only; and (iii) SNPs be independent of other factors which affect the cardioembolic stroke (figure 2). First, we obtained a total of 115 SNPs prominently associated with serum 25(OH)D concentration and served as IVs (P□<□5□×□10^−8^) with linkage disequilibrium (defined as r^2^ = 0.001 and clump distance=10,000 kb). We further withdrew SNPs with horizontal polymorphic effects to persuade the second assumption by searching an online tool (the PhenoScanner database) (21,22). The F-statistic (F□=□beta^2^/se^2^) was obtained to assess the weak IV bias for avoiding weak instrument bias. If the F-statistics>10, the first assumption was satisfied.

**Figure 2:**
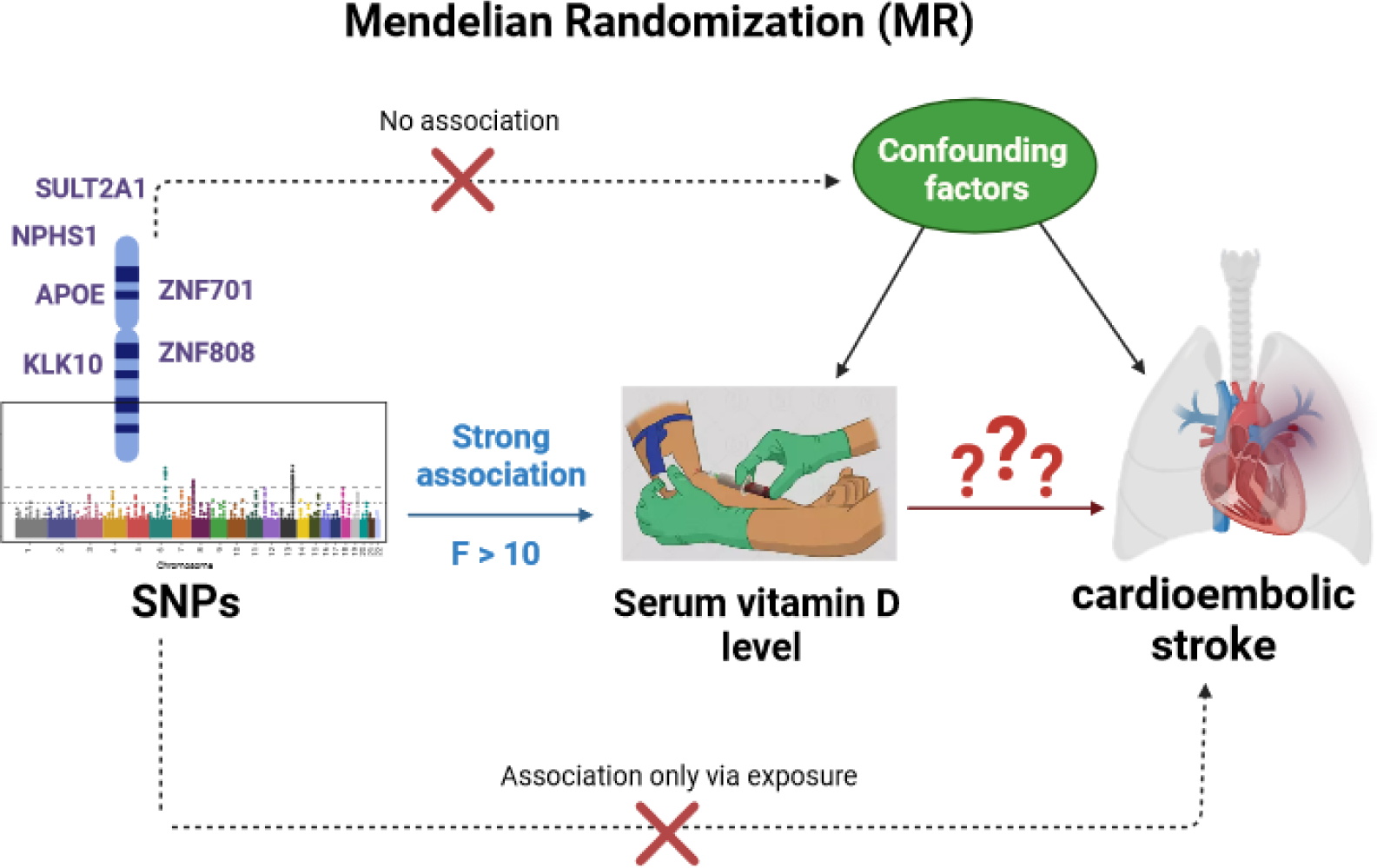
Mendelian randomization design to identify causal risk factor. Exposure is serum 25 hydroxyvitamin D concentration, and outcome is cardioembolic stroke (Created with BioRender.com).

In the harmonization section, the effect allele frequency given in the corresponding GWAS was employed to identify and disregard all palindromic SNPs. The palindromic SNPs with the intermediate allele frequency□>□0.45 and□<□0.55 were excluded from the above-selected SNPs (23). Moreover, SNPs with MAF less than□0.01 were omitted to avoid potential statistical bias from the original GWAS (24).

### Mendelian Randomization Analysis

The primary method was the random-effects multiplicative inverse variance weighted (IVW) to estimate the causal relationship between serum 25(OH)D concentration and cardioembolic stroke (25). Cochran’s Q statistic for MR-inverse-variance weighted analyses used to detect heterogeneity and Rucker’s Q statistic for MR-Egger (26). The I^2^ statistic was also estimated to evaluate the heterogeneity, and the I^2^ values <25%, 25–75%, and >75% were considered to indicate low, moderate, and high heterogeneity, respectively (27). Pleiotropy was assessed using the intercept test with the MR-Egger regression method (28), and horizontal pleiotropy was evaluated with MR-PRESSO (29).

As the IVW method can be affected by pleiotropy or invalid instrument bias (30), we assessed the validity and robustness of the results by executing several sensitivity analyses, including MR-Egger, weighted median, simple mode, and weighted mode method (31). Additionally, we used other approaches, including the Maximum likelihood method, Robust Adjusted Profile Score (RAPS), MR-Lasso, constrained maximum likelihood, mode-based, debiased inverse variance weighted, contamination mixture method and mixture model to reveal the robustness of our results (32–34).

To identify outliers with potential pleiotropy, we used MR-PRESSO and RadialMR (35,36). We also applied Cook’s distance and Studentized residuals to ascertain whether any individual SNPs were detected as outliers and influential points (37). Additionally, Leave-one-SNP-out analysis and its plot performed to assess the influence of potentially pleiotropic SNPs on the causal estimates by leaving each genetic variant out (38). Funnel, forest, and scatter plot depicted to detect directional pleiotropy, to visual association genetic association, and to investigate inspection of outliers and causal estimates, respectively (39).

All statistical analyses were performed using R software (version 4.0.3) by “*TwoSampleMR*,” “*MendelianRandomization*”, “*MR-PRESSO*,” “*RadialMR*”, “*MRMix*”, “*mr.raps*”, and “*GMRP*” packages, and *mrrobust* package STATA (version 17). Results were reported as odds ratios (OR) with corresponding 95%, and associations with P-values between less than 0.05 were regarded as suggestive evidence of associations.

## Results

Application of the selection criteria identified 115 SNPs as possible independent IVs related to serum 25(OH)D concentration after p□<□5□×□10^−8^ and the clumping. In harmonizing, two SNPs removed related to palindromic (rs11606 and rs2246832), and 110 SNPs remained for initial analyzing (Supplementary data). The weak instrumental bias can be statistically ignored, as the F-statistic ranged from 29.78 to 2567.54 (Supplementary data).

At first, the estimation accuracy did not satisfy, and we attempted to enhance it by identifying potential outliers through the MR-PRESSO, RadialMR, Cook’s distance, and Studentized residuals. Among them, Cook’s distance and Studentized residuals outperformed others, as shown figure 3.

**Figure 3:**
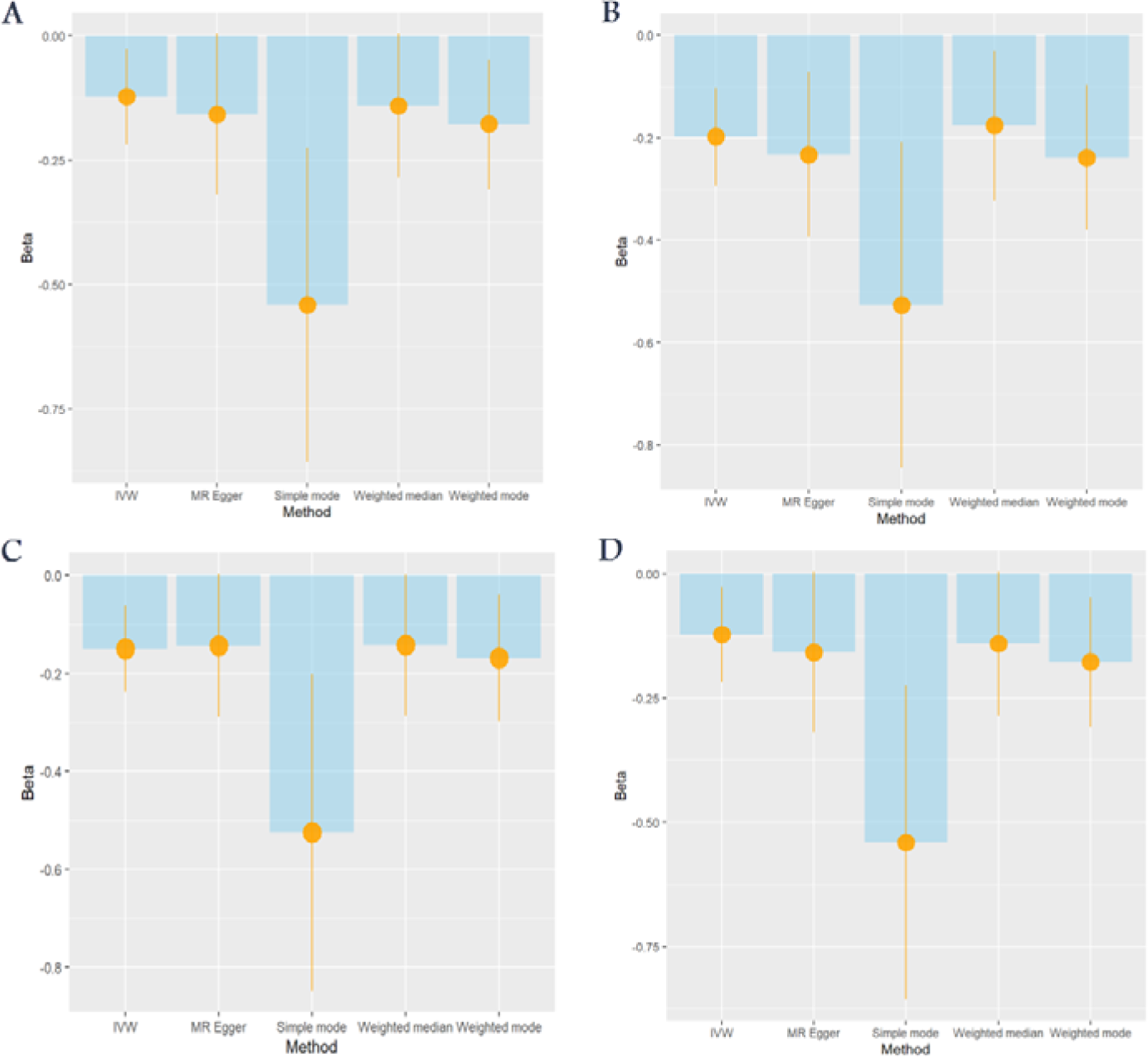
Initialing Mendelian randomization. A: results with potential outlier and influential points, B: results after using Cook’s distance and Studentized residuals. C: results after using Radial-MR. D: results after using MP-PRESSO. The height of each column indicates the beta along with the standard error, and the horizontal axis indicates the types of methods. Among them, Cook’s distance and Studentized residuals outperformed others.

To roughly display relationship of the undefined causal variables to cardioembolic stroke of study, we used beta of serum 25(OH)D concentration regressed on the undefined causal variable versus the cardioembolic stroke. Regression coefficients revealed positive association between serum 25(OH)D concentration and cardioembolic stroke, as shown in figure 4 and Supplementary Figure S2. Additionally, the chromosomal histogram of selected SNPs and their position annotation has been exhibited in Supplementary Figure S3.

**Figure 4:**
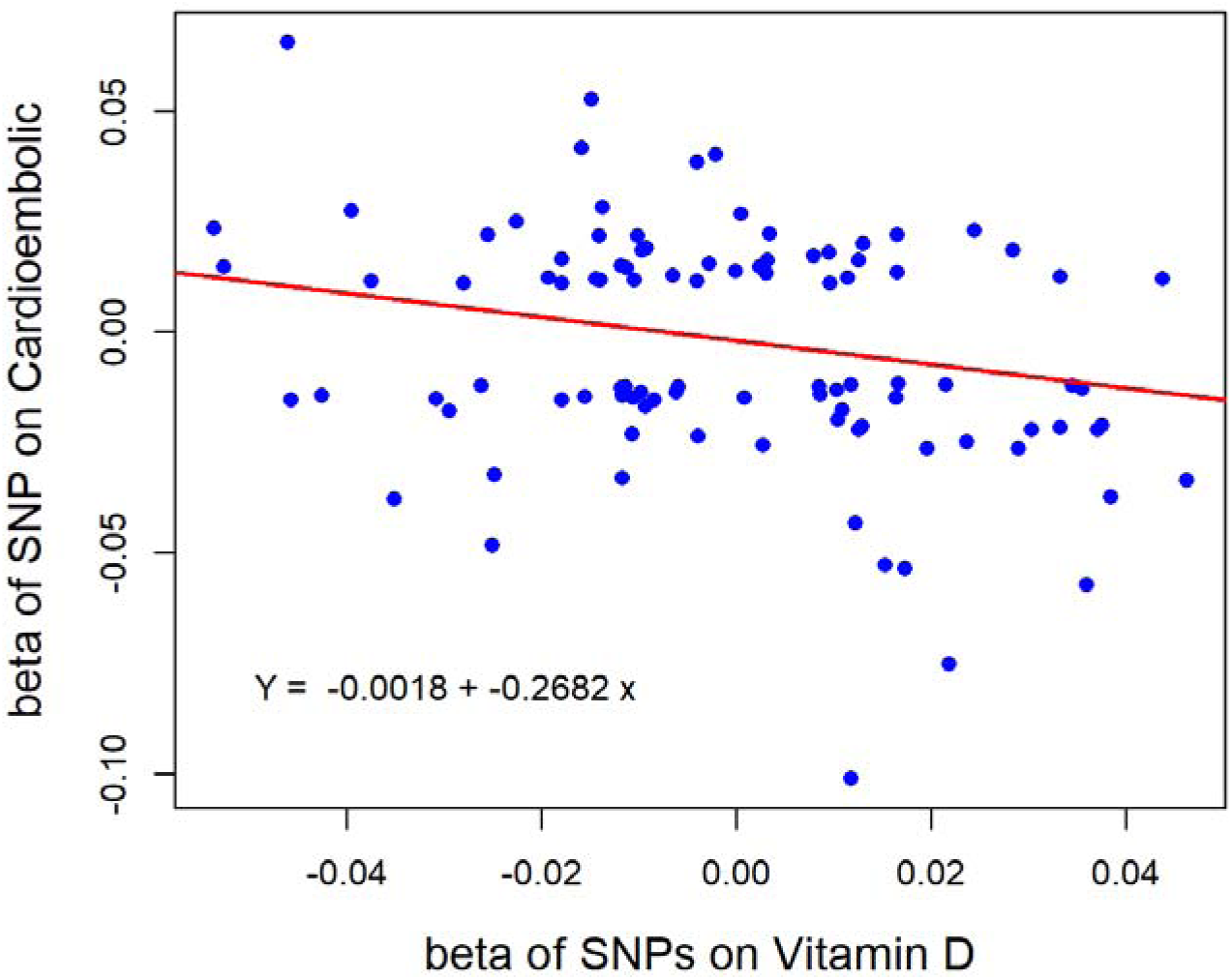
Two-way Scatter plots for beta values of disease and undefined causal variables.

We identified (with N=99 SNPs) a causal relationship between serum 25(OH)D concentration and cardioembolic stroke using IVW [OR□=□0.82, 95% CI: 0.67-0.98, p□=□0.037]. Horizontal pleiotropy was not seen by the MR-Egger intercept-based test [MR-Egger intercept□=□0.001; p□=□0.792], suggesting an absence of horizontal pleiotropy. Cochrane’s Q value [Q=78.71, p-value□=□0.924], Rucker’s Q [Q=78.64, p-value=0.913], and I^2^=0.0% (95% CI: 0.0%, 24.6%) statistic suggested no heterogeneity in the connection between serum 25(OH)D concentration and cardioembolic stroke.

This finding remained consistent using different Mendelian randomization methods, including Maximum likelihood [OR=0.82, 95%CI: 0.67-0.98, p-value=0.036], Constrained maximum likelihood method [OR=0.76, 95%CI: 0.64-0.90, p-value=0.002], Debiased inverse-variance weighted method [OR=0.82, 95%CI: 0.68-0.99, p-value=0.002], MR-PRESSO [OR=0.82, 95%CI 0.77-0.87, p-value=0.022], RAPS [OR=0.82, 95%CI 0.67-0.98, p-value=0.038], MR-Lasso [OR=0.82, 95%CI 0.68-0.99, p-value=0.037], but were not significant for MR-Egger, Median, MRMix (Supplementary Figure S4) and mode (Supplementary Figure S5) methods in Figure 5. The readers can find a step-by-step demonstration of all results by visiting the GitHub: https://akbarzadehms.github.io/VitDcardioembolicStrokeMR/.

**Figure 5:**
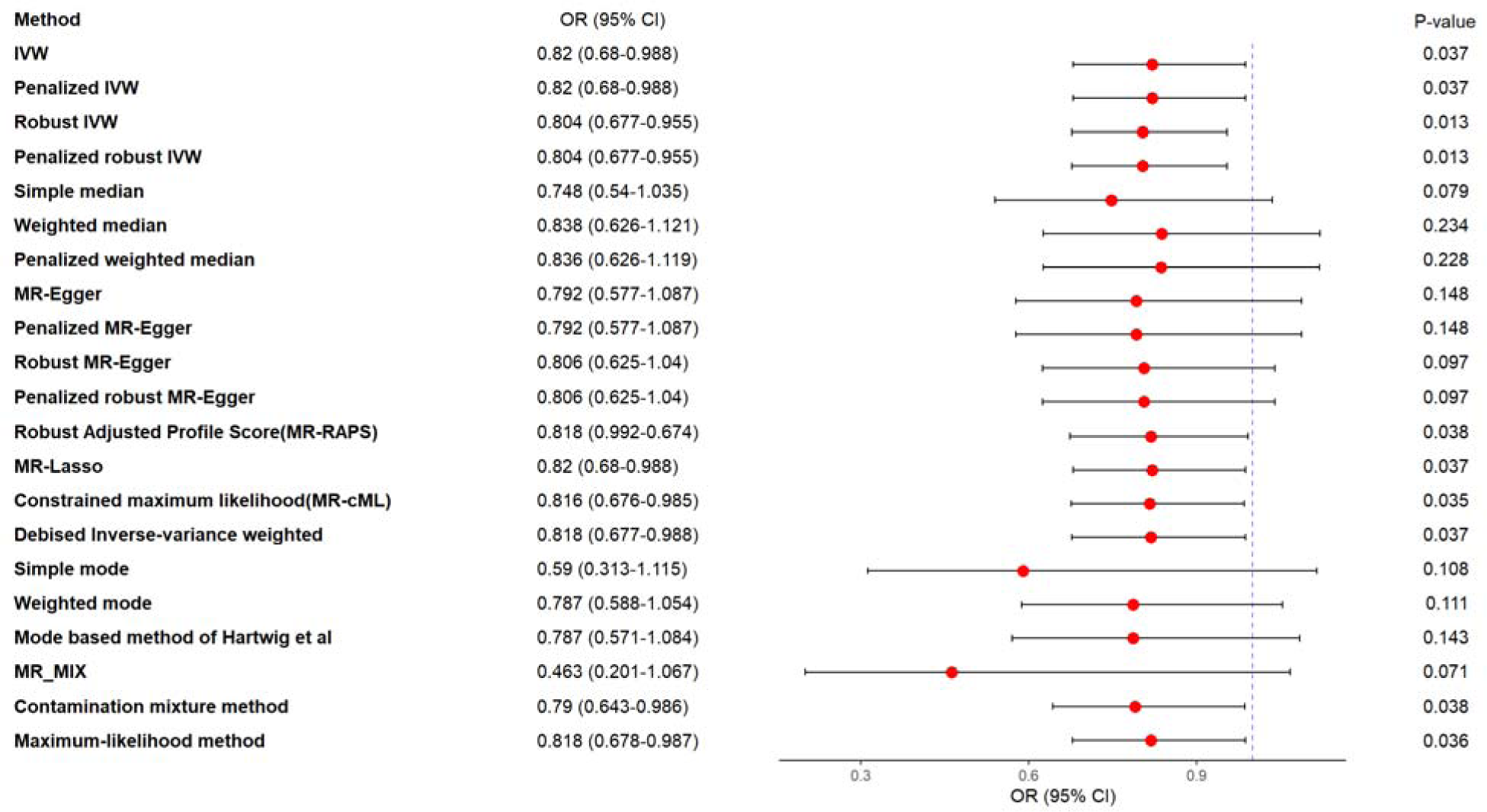
Mendelian randomization analysis according to several methods. Results were exhibited based on the odds ratio (OR) and its confidence interval of 95%. All methods reveal OR<1. Among them, Inverse variance weighted, Maximum likelihood, constrained maximum likelihood, Debiased inverse-variance weighted, Robust Adjusted Profile Score, and MR-Lasso methods were significant. MR-Mix, MR-Egger, Median, and mode methods indicated a suggestive inverse association.

### Sensitivity Analyses

According to the results of Cochran’s Q test, there was not statistically significant heterogeneity observed. MR-Egger regression intercept analysis indicated that there was no directional horizontal pleiotropy in cardioembolic stroke (Supplementary Figure S6). MR pleiotropy residual sum and outlier test (MR-PRESSO) methods, MR pleiotropy residual sum and outlier (Radial MR), Cook’s distance, and Studentized residuals were used to assess, the potential the outlying SNPs (Supplementary Figure S7). Additionally, we performed the visual examination of scatter plots (Supplementary Figure S8) and leave-one-out plots (Supplementary Figure S9) to assess the influence of outlying values. Funnel and forest plot of causal association depicted for serum 25(OH)D concentration on cardioembolic stroke (Supplementary Figure S9 and S10). Weak instrumental bias was checked via the F-statistic (Supplementary data). Additionally, we assessed whether SNPs within the HLA region was available in our MR analysis (Supplementary data). MR Steiger test also demonstrated that correct causal direction was existed for cardioembolic stroke (P_MR-Steiger_ <0.05).

## Discussion

The results of this study support a causal relationship between a genetically determined change in 25(OH)D levels and the risk of cardioembolic stroke. In this MR study, we discovered an inverse association between genetically determined 25(OH)D and risk of cardioembolic stroke. The odds ratio for cardioembolic stroke was 0.82 per unit increase in log-transformed 25(OH)D. This effect was statistically significant, with a 95% confidence interval of 0.67 - 0.98 and p-value of 0.037. These findings are consistent with previous observational studies that have shown an association between low serum levels of 25(OH)D and recurrent stroke (40,41). An observational study which investigated the association of serum 25(OH)D deficiency with ischemic stroke and subtypes in Indian patients, reported that 25(OH)D deficiency was prevalent. And among the stroke patients, the deficiency was most common in those with large artery atherosclerosis (54.9%), followed by CES (54%) (42). This study revealed that 25(OH)D deficiency had an independent association with ischemic stroke. The association was established in large artery arthrosclerosis and cardioembolic stroke (42). Moreover, A systematic review and meta-analysis synthesized data from nineteen studies and demonstrated that lower vitamin D status is associated with an increased risk of stroke. The pooled relative risk was 1.60 (95% CI: 1.33– 1.92) in this study (12).

In the present study, we conducted a MR analysis to explore the potential causal relationship between 25(OH)D and CES. We utilized a large dataset derived from a genome-wide association study involving individuals of European ancestry. Our results contradict those of a recent MR study based on the extracted summary-level data from the MEGASTROKE consortium for ischemic stroke among individuals of European descent. This study assessed the links between serum 25(OH)D concentrations and various subtypes of ischemic stroke and failed to provide evidence supporting a causal link between higher serum 25(OH)D concentrations within the normal range and ischemic stroke or its subtypes (43). This previous investigation reported that a genetically predicted one standard deviation increase (1-SD) in serum 25(OH)D concentrations did not exhibit a significant association with overall ischemic stroke (OR, 1.01; 95%, CI, 0.94– 1.08; P=0.84). Furthermore, an MR study investigating the association between 25(OH)D and ischemic stroke utilized cohort sample data for genetic analysis, resulting in smaller datasets. For instance, one study reported an odds ratio of 0.64 (95% CI, 0.42–0.91) for the causal role of 25(OH)D in recurrent or de novo ischemic stroke (44).

Another study, using the MR approach in 116,655 individuals from the general population, revealed that genetically low 25(OH)D concentrations were associated with hypertension as an important risk factor for ischemic stroke. However, due to overlapping confidence intervals, they could not conclude a causal relationship between genetically lowered 25(OH)D concentration and ischemic stroke (45). Another MR study on subtypes of ischemic stroke indicates that there is no significant causal relationship between 25(OH)D concentration and various cerebral small vessel disease (cSVD)-related conditions, including lacunar stroke, white matter hyperintensity, cerebral microbleeds, and white matter, basal ganglia, and hippocampal perivascular spaces. The study’s results remained consistent even when considering genetic factors. However, the research did reveal a negative association between cerebral microbleeds and 25(OH)D concentration (46).

While vitamin D is primarily known for its role in regulating calcium absorption and bone health, it also has important functions in various other tissues and cells throughout the body, including vascular smooth muscle cells, platelets, and immune cells(15). Since important role of these cells in the pathogenesis of ischemic stroke, they may be a possible mechanism that links vitamin D and stroke. Several explanatory mechanisms may elucidate cardiovascular benefits of 25(OH)D. Serum 25(OH)D holds direct protective vascular effects via increasing epithelial progenitor cells (EPCs), and improving glycemic control (13). Additionally, 25(OH)D possesses diverse paracrine and autocrine properties that modulate the immune system. For example, it downregulates the production of proinflammatory cytokines, including tumor necrosis factor-α and IL-6 (47). A clinical trial conducted on ischemic stroke patients has indicated that 25(OH)D could ameliorate TLR4/NF-kβ signaling pathway after ischemia, as this pathway and its downstream proinflammatory cytokines IL-1β and IL-6 expression were found elevated in ischemic stroke patients(48). Moreover, 25(OH)D may contribute in moderating thrombogenesis and atherosclerotic processes, mainly through modulating free radical formation and atherosclerotic inflammatory cytokines release (49).

MR analysis functions as an alternative approach to that of to a randomized controlled trial (50). It would be impractical and somewhat unethical to randomize vitamin D deficiency. However, MR analysis offers a viable approach to explore the potential causal impact of low 25(OH)D concentration on cardioembolic stroke. Additionally, MR analysis is capable of estimating the long-term effects of an exposure, a task that would be unattainable in a randomized clinical trial. Consequently, MR analysis is a valuable option for addressing the question of whether a vitamin D supplement could offer benefits in preventing CES. The MR approach implemented in this study has some advantages in strengthening causal inference. Its design decreases potential confounding or reverse causation which are present in observational studies. Furthermore, we did not detect any violations of the assumptions of Mendelian randomization to the extent that they could be tested, and our instruments used for instrumental variable analyses were strong with an F-value ranged from 29.8 to 2567.5.

Our study has several limitations. The effect size suggests a moderately protective effect of increased 25(OH)D on cardioembolic stroke. Although the implication of a positive finding in a Mendelian randomization study is the presence of causality, it’s essential to bear in mind that this setting differs from a randomized intervention trial. In our investigation, we examined the effects of lifelong exposure to low 25(OH)D levels rather than short-term interventions with vitamin D. Furthermore, since the GWAS employed in our research predominantly involved patients of European ancestry, the applicability of our findings to other ethnic groups could be restricted. We were unable to address the non-linear association in the analysis, as this would require a single dataset containing all exposure and outcome variables, along with detailed information on the genetic instruments.

Overall, while our MR findings provide compelling evidence for a causal relationship between higher 25(OH)D levels and low CES rate. Few MR studies have specifically examined cardioembolic stroke, and our study power is limited. Larger GWAS of ischemic stroke subtypes would enhance power for MR analyses. Further research should explore potential threshold effects.

## Conclusion

In summary, our MR analysis provides suggestive evidence that increased 25-Hydroxyvitamin D levels may play a causally protective role in the development of cardioembolic stroke. Additional MR studies in larger samples could help clarify this relationship and elucidate the magnitude of effect. Determining the role of 25-Hydroxyvitamin D in stroke subtypes has important clinical and public health implications. Further research integrating genetic epidemiology, molecular pathways, and clinical data is warranted to gain a more definitive understanding of the connections between vitamin D biology and CES.

## Supporting information

Supplementary Dataset: Serum 25 hydroxyvitamin D concentration on cardioembolic stroke

## Declarations

## Acknowledgments

We want to acknowledge the participants and investigators who made summary data available.

## Consent for publication

Not applicable.

## Data Availability Statement

The original data used are publicly available at https://gwas.mrcieu.ac.uk/. The original contributions presented in the study are included in the article/Supplementary Material; The process of MR analyses and the results are publicly available through the following HTML link: https://akbarzadehms.github.io/VitDcardioembolicStrokeMR/

## Funding

Not applicable.

## Ethical approval and consent to participate

The Isfahan University of Medical Sciences ethics committee approved this study (Research Approval Code: 340111 & Research Ethical Code: IR.MUI.RESEARCH.REC.1401.063). Informed consent was obtained from all subjects in the original genome-wide association studies. The research has been performed by the Declaration of Helsinki.

## Author Contributions

DH, and MA conceived the study, analyzed the data, and drafted the initial manuscript. LNHB generated Tables and Figures. FH, NE, and AHS investigated. FH, NE, AST, AHS, HH, FA, MH, MSD, and MM, revised the manuscript. MA and MM supervised, edited, and finalized the manuscript. All authors reviewed and approved the final manuscript.

## Conflict of Interest

All authors declare that they have no conflicts of interest.

## Supplementary materials

**Figure S1.**
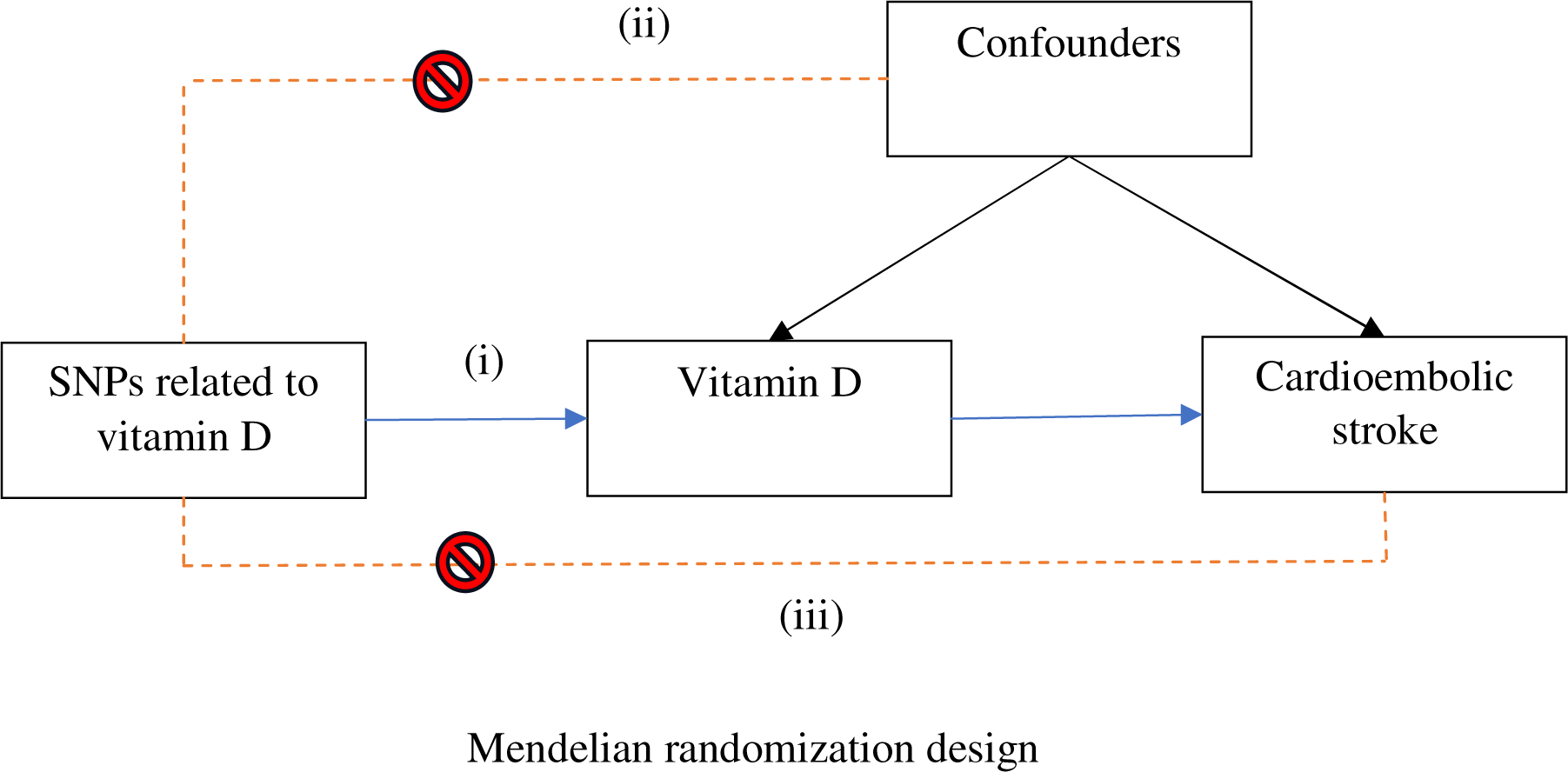
Diagram of Mendelian randomization framework in the current paper. The instrumental variable (IV) techniques are a few available ways to estimate the causal effects without the full knowledge of all the confounders of the exposure-outcome association. The IV is referred to as an external variable (SNPs) that is associated with exposure. Besides, it is independent of the outcome and any factor linked to it, other than exposure. MR utilizes genetic variation as an instrumental variable (IV) to investigate the causal association between exposure and outcome in non-experimental data (1). To utilize a genetic variant to be a valid instrumental variable, several main assumptions should be satisfied. The IVs (SNPs) are strongly associated with exposure (s) and should be clear quantifiably. (ii) The SNPs are not linked with any confounder of the exposure-outcome association. (iii) The SNPs do not affect the outcome, except possibly through their association with the exposure (s).

**Figure S2.**
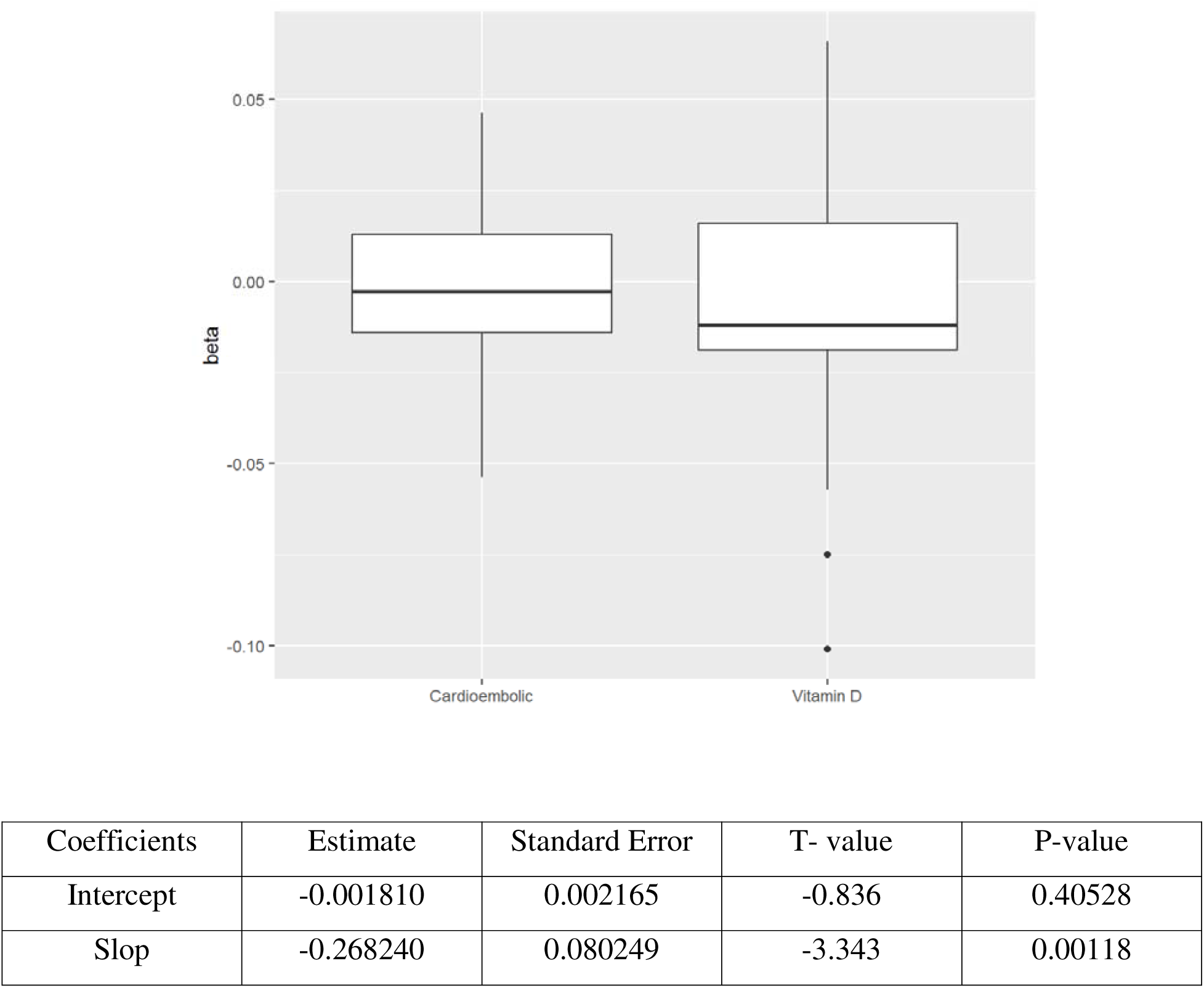
Display relationship of the undefined causal variables between serum 25 hydroxyvitamin D concentration and cardioembolic stroke.

**Figure S3.**
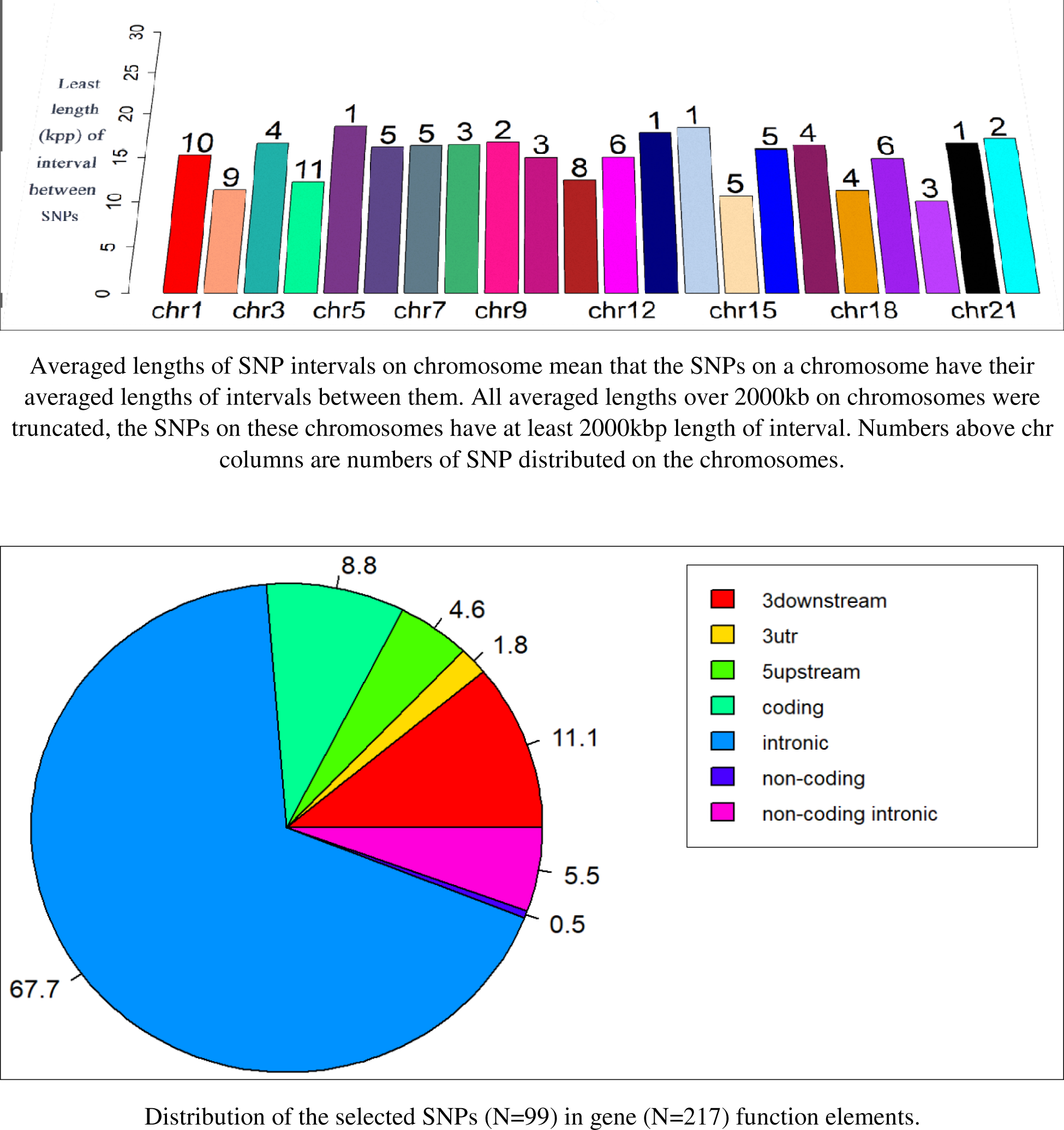
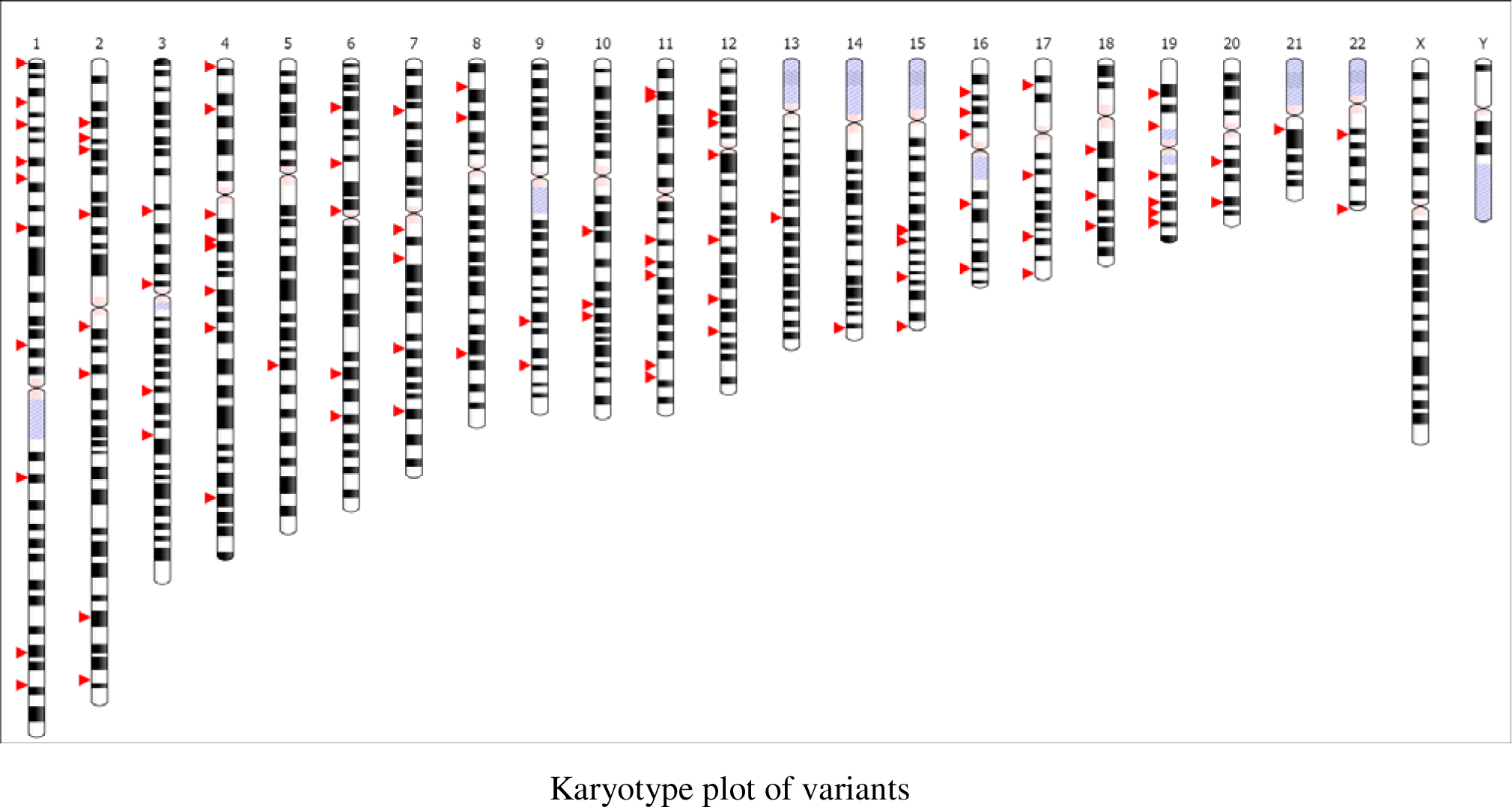
SNP Position Annotation. For this purpose, we used position annotation analysis of □selected SNP□s (chromosome number and SNP position of hg19). Position annotation analysis provides position information of these selected SNPs on chromosomes such as chromosome distribution and averaged interval between SNPs. Therefore, as the following shows, we depicted a histogram for averaged distances between SNPs and SNP numbers on chromosomes.

**Figure S4.**
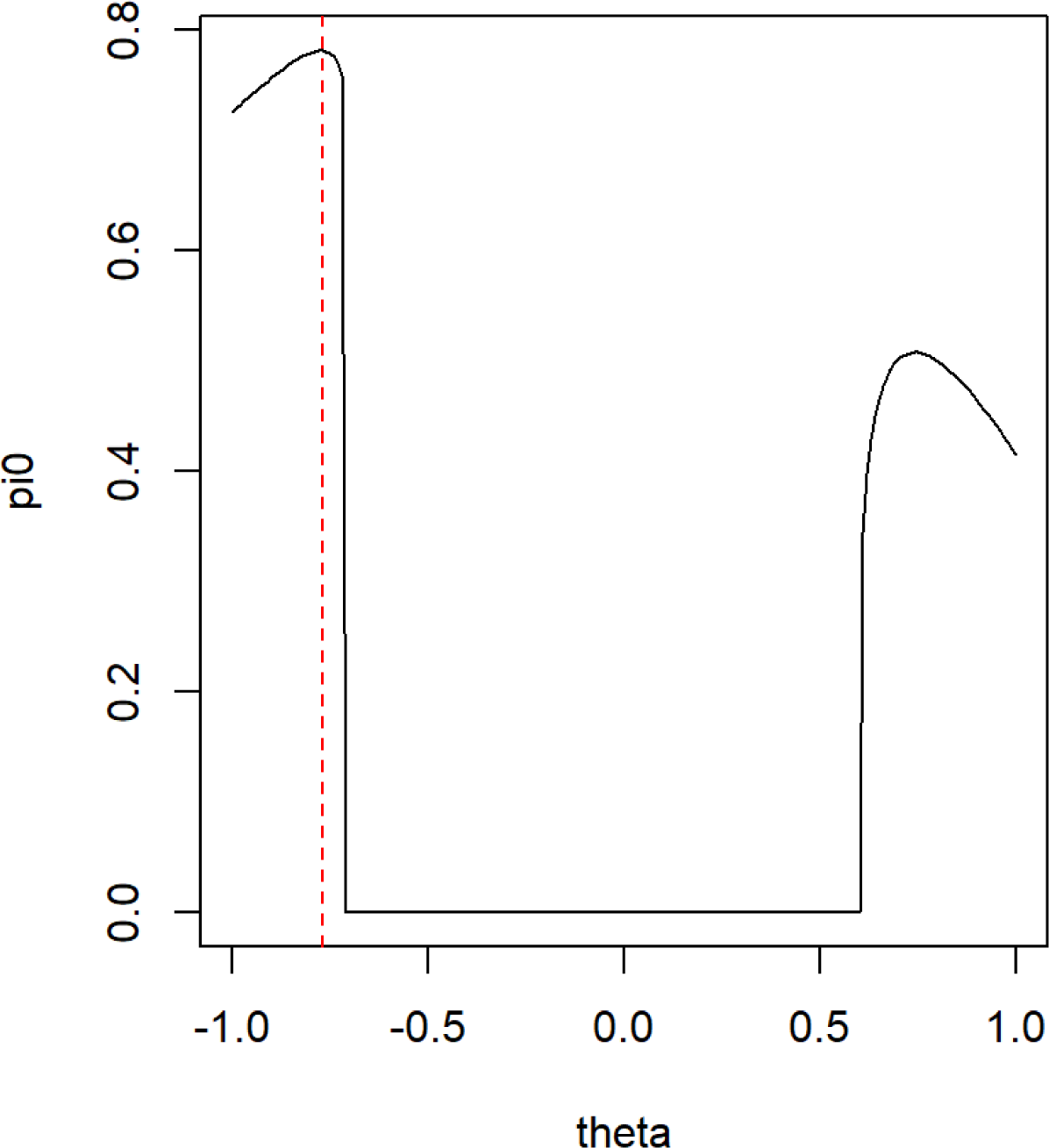
Profile plot. Plot theta (estimate of causal effect) against pi0 (the probability mass of the null component corresponding to the estimated theta). Profile plot made of smooth curve with a clear maximum indicates stable performance. For each fixed theta, it fits the following mixture model

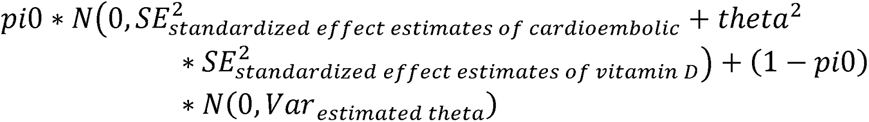 on the residual of (standardized effect estimates of cardioembolic* standardized effect estimates vitamin D). Then, it selects the value of theta that leads to the maximum pi0 as the estimate of causal effect [OR=0.46, 95%CI 0.19-1.09, p-value=0.071].

**Figure S5.**
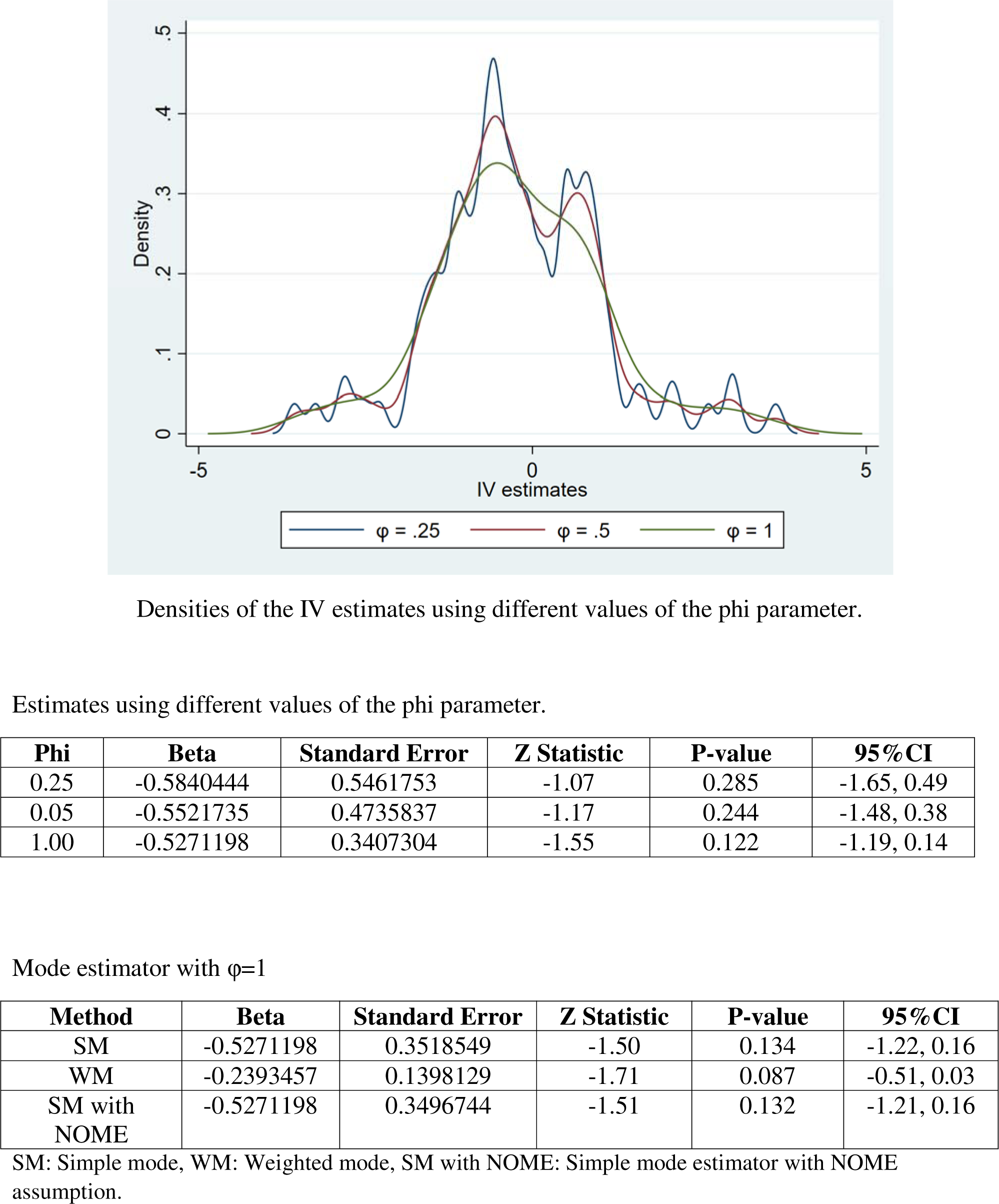
Densities of the IV estimates using different values of the phi parameter.

**Figure S6.**
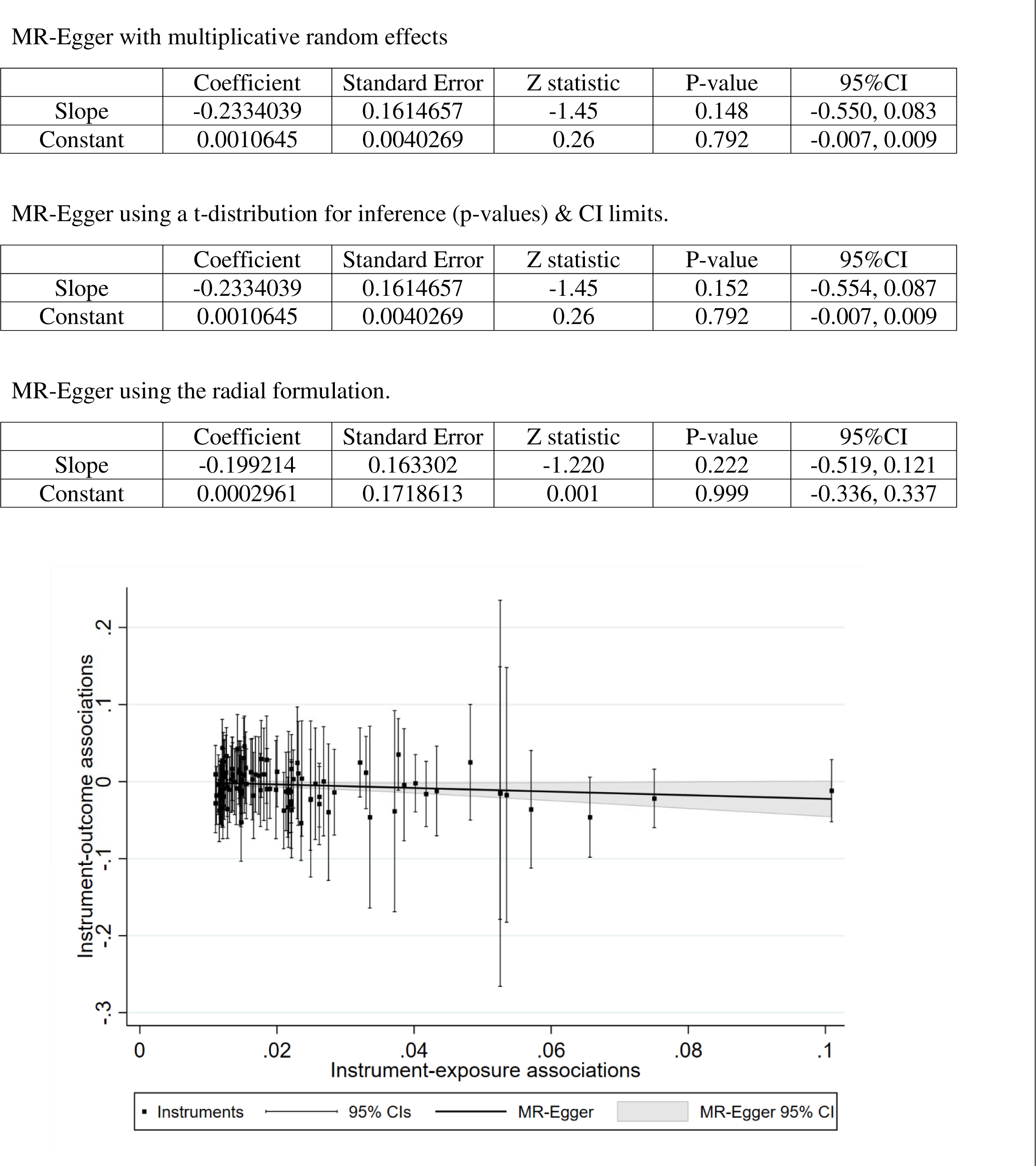
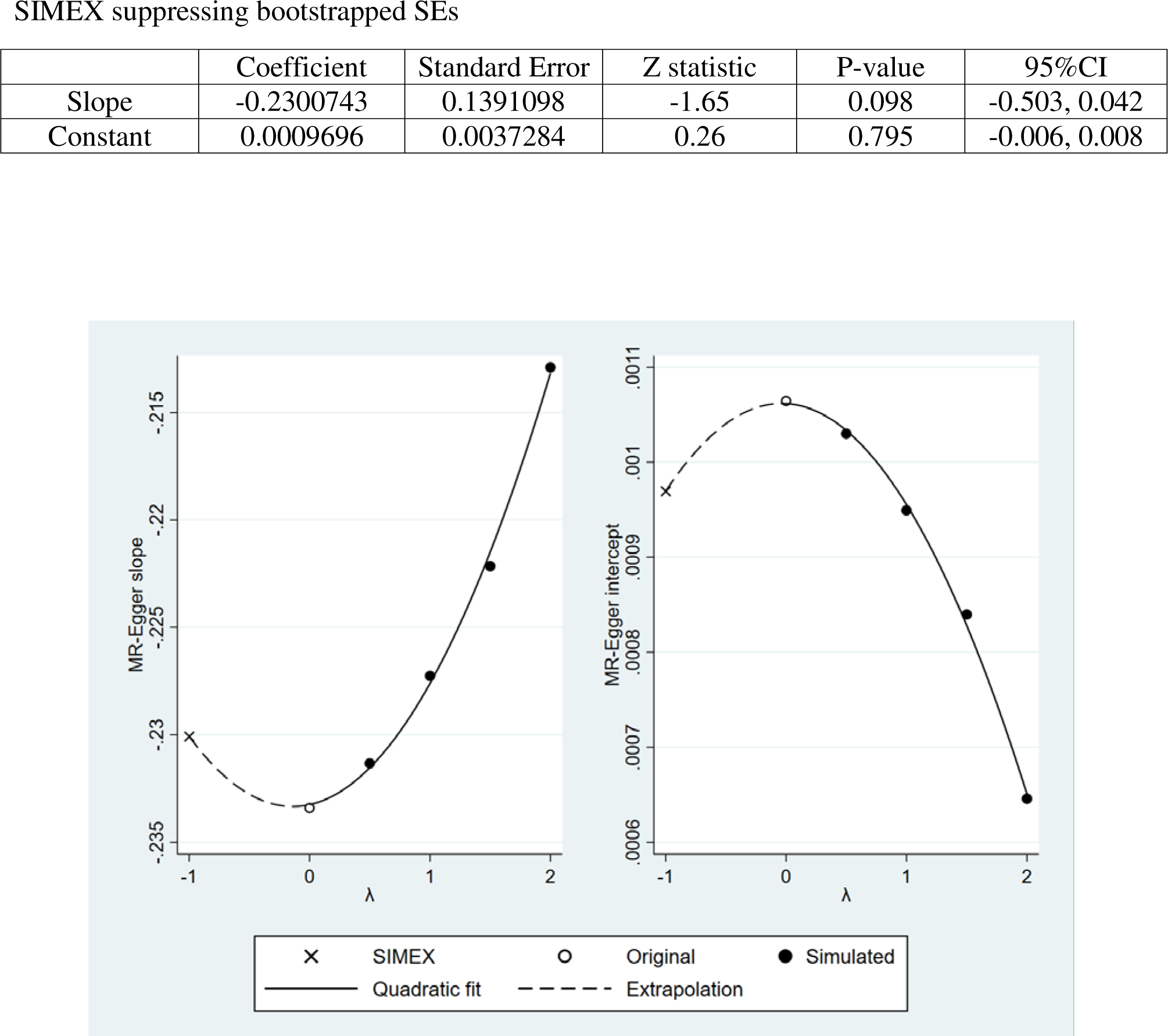
Scatter plot showing the genotype summary level data points and the fitted MR-Egger model.

**Figure S7.**
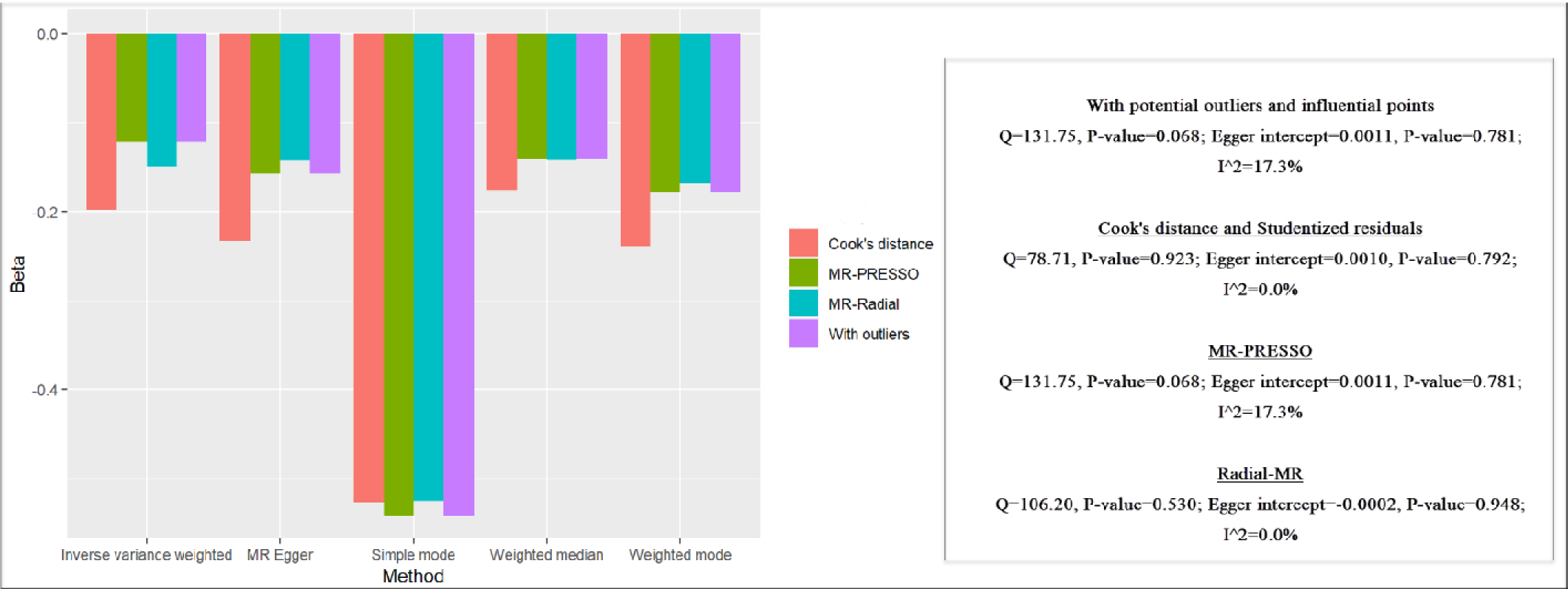
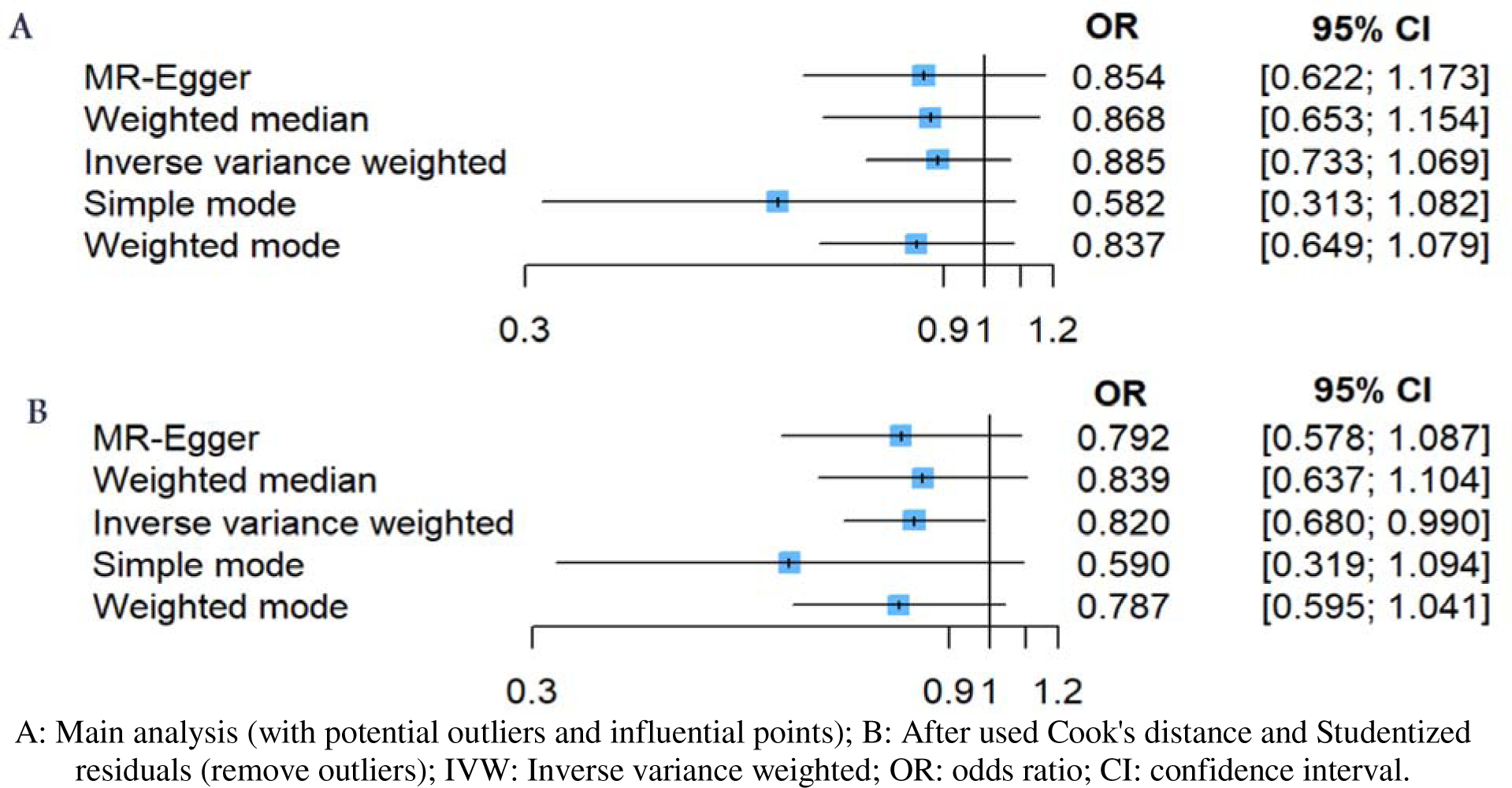
The investigating of outliers based on MR-PRESSO, RadialMR, and Cook’ distance and Studentized residuals. To identify potential outliers and influential SNPs, we utilized several methods, including MR-PRESSO, RadialMR, and Cook’s distance and Studentized residuals. The following figure shows the results of them. As seen, Cook’s distance was outperformed by others. As a result, we used Cook’s distance to detect the potential outlier and influential SNPs. Primary analysis showed that the estimation accuracy did not satisfy [Q=131.75, P-value= 0.068; I^2^=17.3%; Egger intercept= 0.0011, P-value=0.781]. After applying MR-PRESSO [Q=131.75, P-value= 0.068; I^2^=17.3%; Egger intercept= 0.0011, P-value=0.781], RadialMR [Q=106.20, P-value=0.530; I^2^=0.0%; Egger intercept=-0.0002, P-value=0.948], and Cook’s distance [Q=78.71, P-value=0.923; I^2^=0.0%; Egger intercept=-0.0010, P-value=0.792], we found that Cook’s distance and Studentized residuals outperformed others.

**Figure S8:**
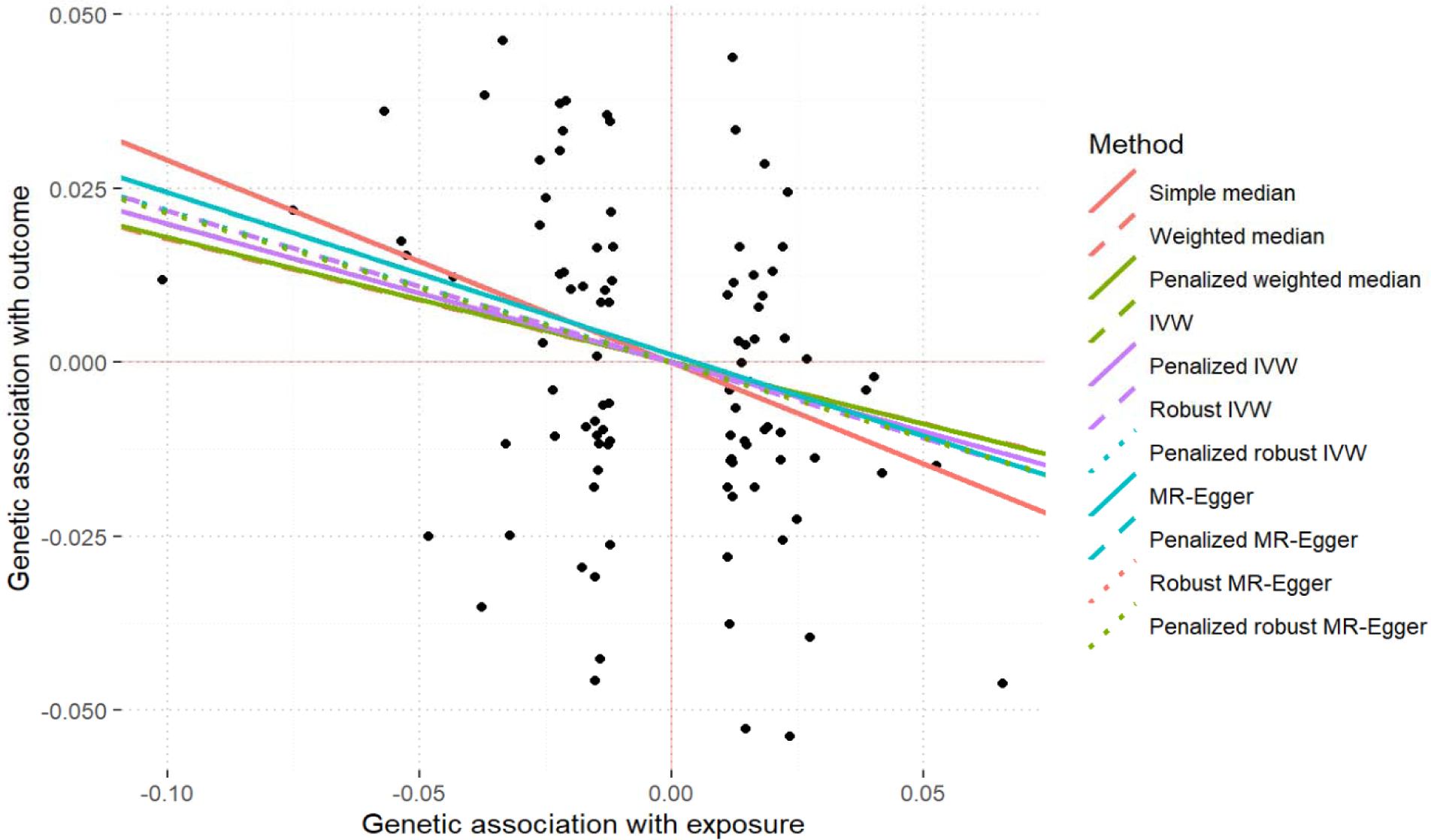
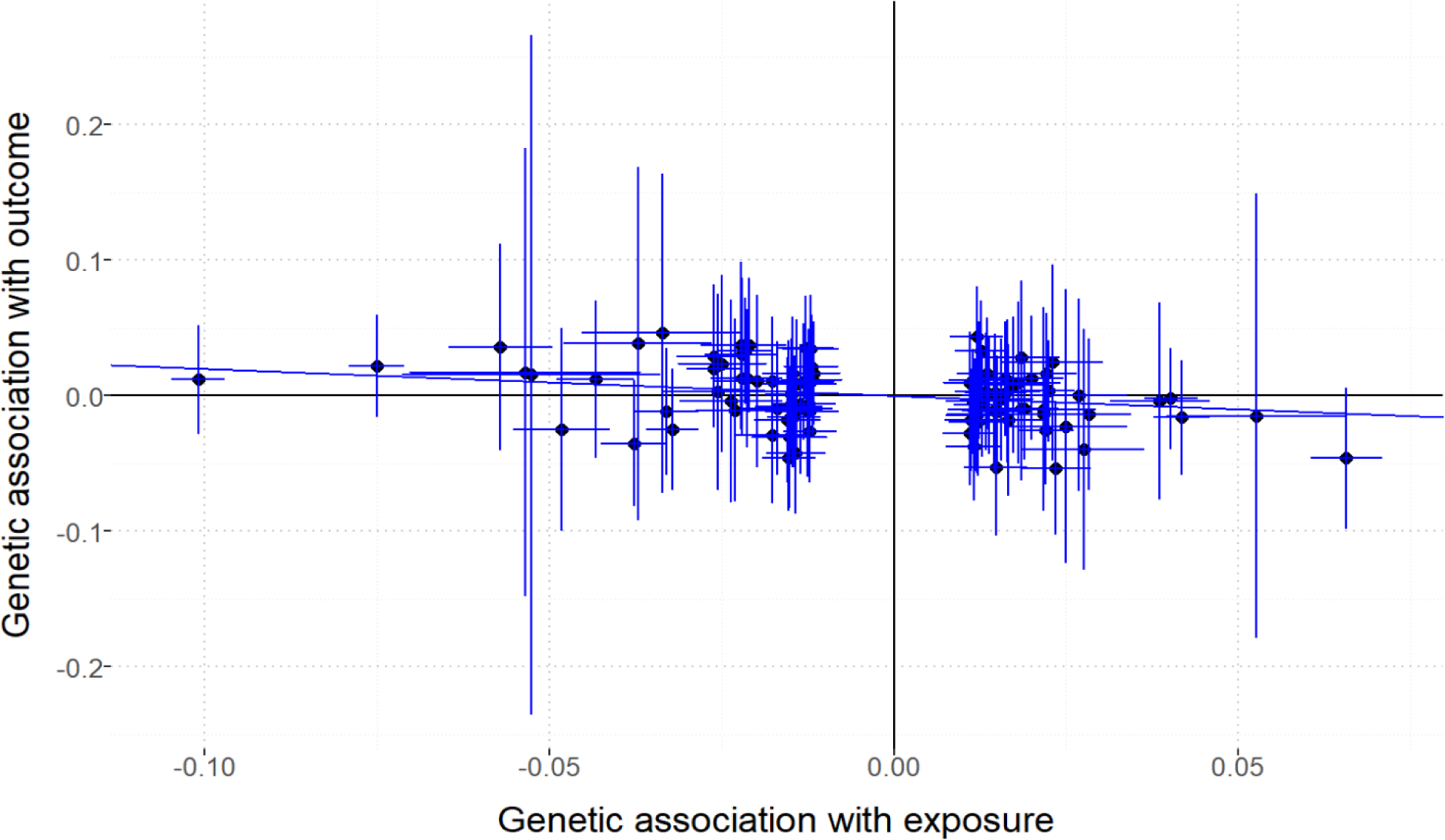
Comparison of the causal estimates from the various Mendelian randomization methods and scatter plot of the potential effects of serum 25 hydroxyvitamin D concentration and cardioembolic stroke. The following figures show the several Mendelian randomization methods, including Simple median, Weighted median, Penalized weighted median, Inverse variance weighted, Penalized Inverse variance weighted, Robust inverse variance weighted, Penalized robust inverse variance weighted, MR-Egger, Penalized MR-Egger, Robust MR-Egger, Penalized robust MR-Egger, Maximum likelihood estimate, and Mode-based method. All methods elucidate negative association between of serum 25 hydroxyvitamin D concentration and cardioembolic stroke. The following figure displays a scatter plot, which shows a genetic association. X-axis lies with the genetic association of exposure (serum 25 hydroxyvitamin D concentration), and the y-axis is the genetic association of outcome (cardioembolic stroke). The linear line corresponds to the Inverse variance weighted method. As seen, slop is negative.

**Figure S9:**
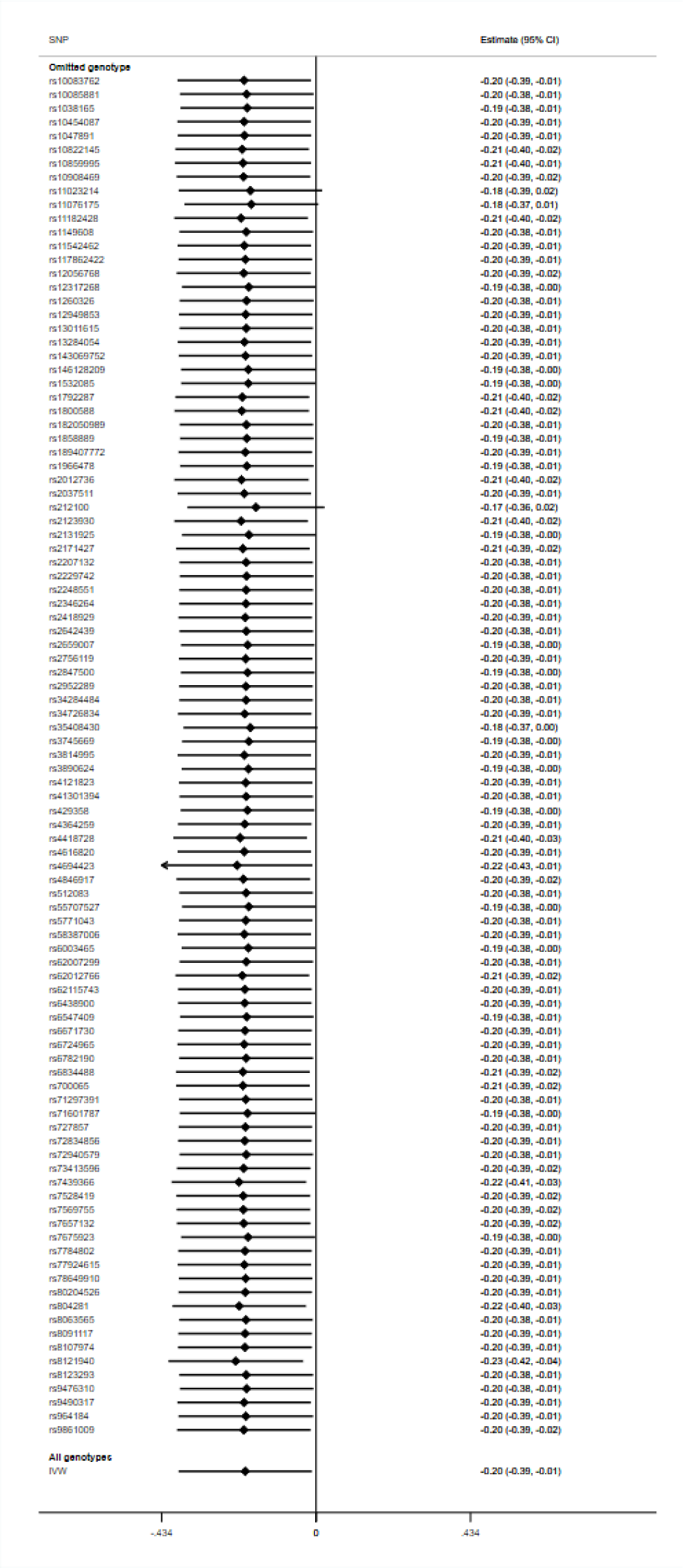
Leave-one-out plot to assess if a single variant is driving the association between serum 25 hydroxyvitamin D concentration and cardioembolic stroke.

**Figure S10.**
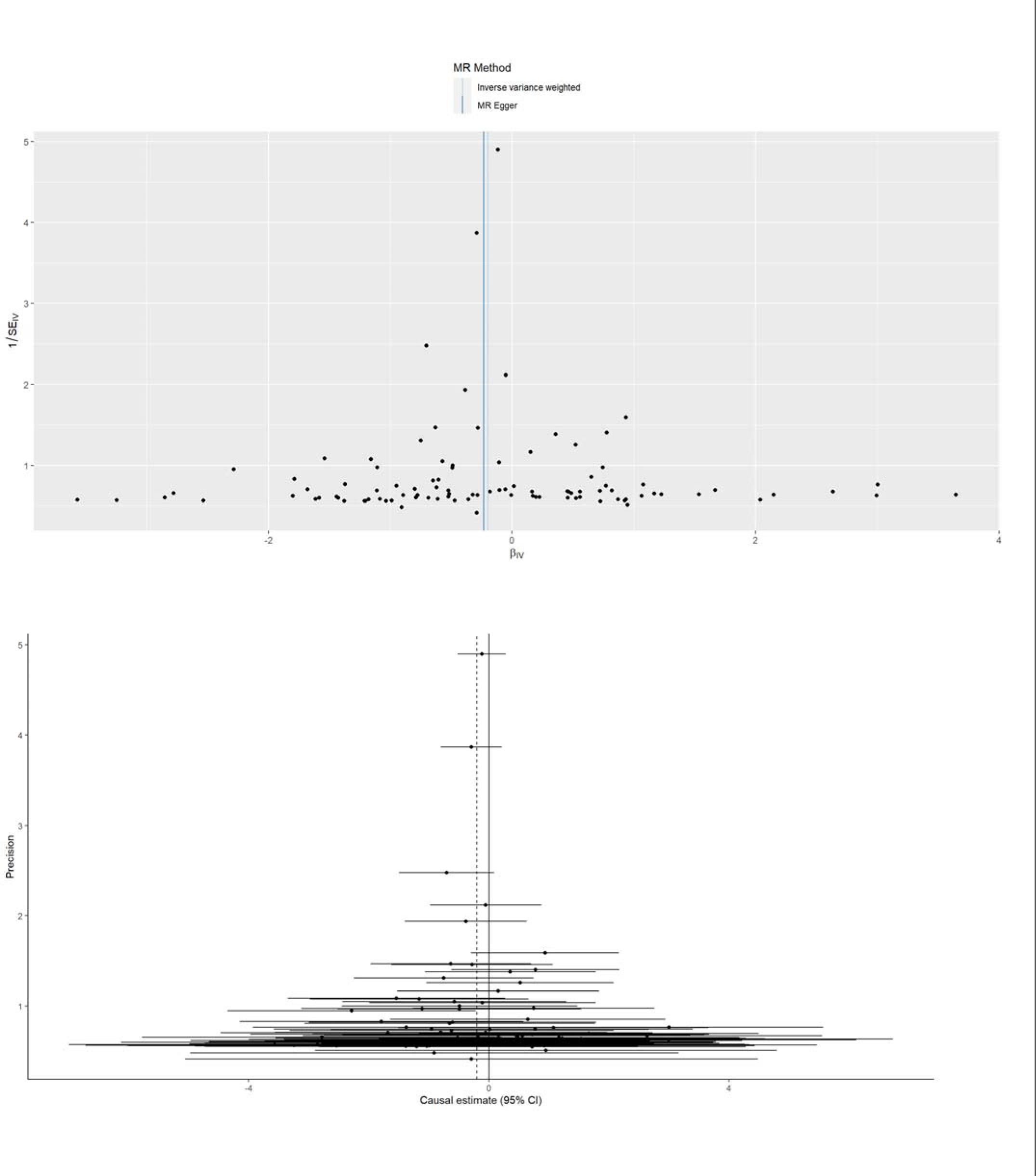
Funnel plot of causal association between serum 25 hydroxyvitamin D concentration and cardioembolic stroke.

**Figure S11:**
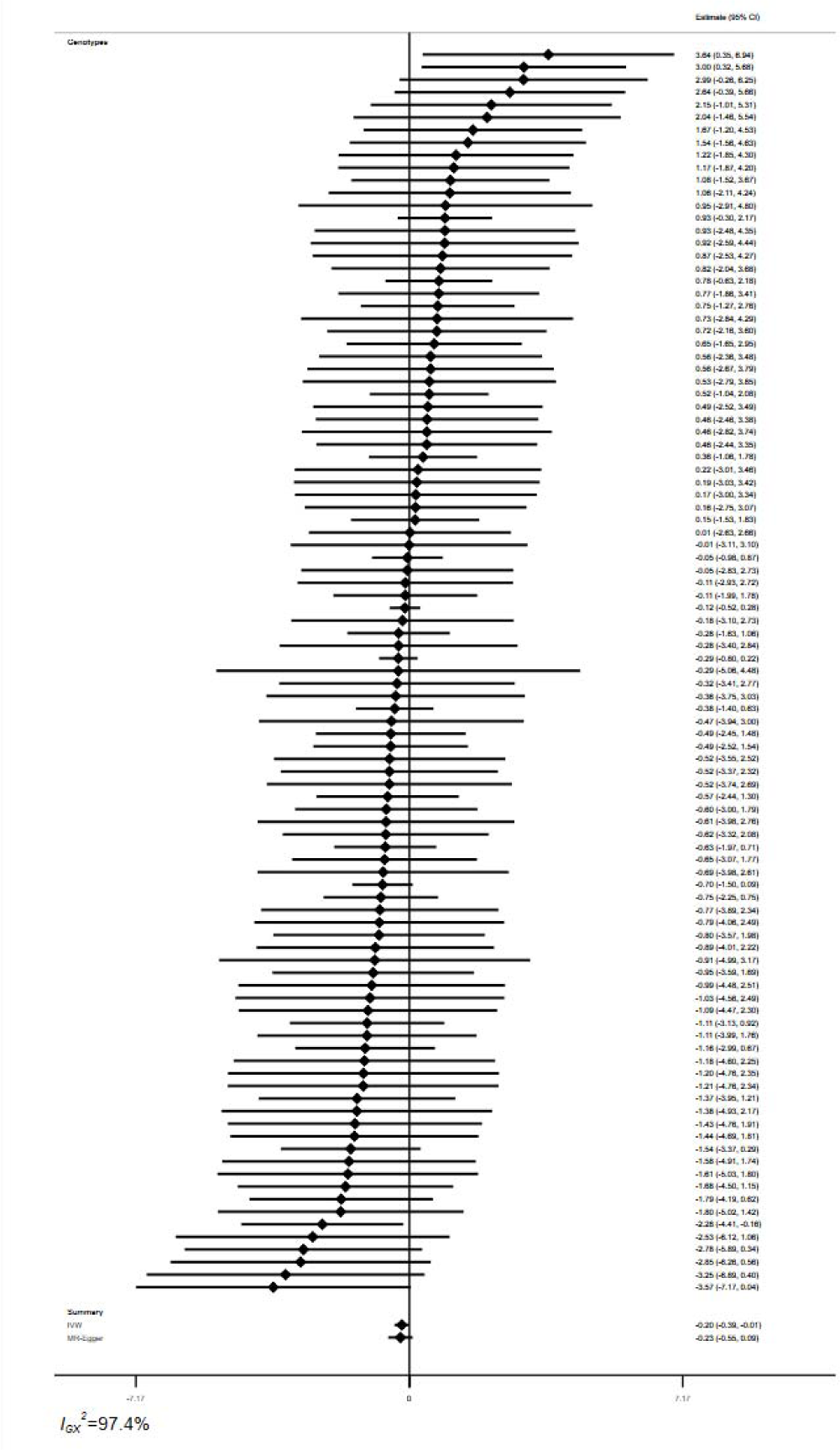
Forest plot of variant specific inverse variance estimates for causal association between serum 25 hydroxyvitamin D concentration and cardioembolic stroke.

## Notes

### Competing Interest Statement

The authors have declared no competing interest.

### Funding Statement

This study did not receive any funding.

### Author Declarations

The dataset employed in this study was obtained from the GWAS catalog (https://www.ebi.ac.uk/gwas/). for vitamin D measurement the study accession is GCST90000614 and for cardioembolic stroke the study accession is GCST90104541.

## References

1. Krishnamurthi R V, Feigin VL, Forouzanfar MH, Mensah GA, Connor M, Bennett DA, et al. Global and regional burden of first-ever ischaemic and haemorrhagic stroke during 1990–2010: findings from the Global Burden of Disease Study 2010. Lancet Glob Heal. 2013;1(5):e259–81.

2. Thrift AG, Howard G, Cadilhac DA, Howard VJ, Rothwell PM, Thayabaranathan T, et al. Global stroke statistics: An update of mortality data from countries using a broad code of “cerebrovascular diseases.” Int J Stroke. 2017;12(8):796–801.

3. Long X, Lou Y, Gu H, Guo X, Wang T, Zhu Y, et al. Mortality, recurrence, and dependency rates are higher after acute ischemic stroke in elderly patients with diabetes compared to younger patients. Front Aging Neurosci. 2016;8:142.

4. Fatisa Y. Reference 5. Vol. 10, Jurnal Peternakan Vol Februari. 2013. p. 31–8.

5. Adams Jr HP, Bendixen BH, Kappelle □, Biller J, Love BB, Gordon DL, et al. Classification of subtype of acute ischemic stroke. Definitions for use in a multicenter clinical trial. TOAST. Trial of Org 10172 in Acute Stroke Treatment. stroke. 1993;24(1):35–41.

6. Mohr JP, Caplan LR, Melski JW, Goldstein RJ, Duncan GW, Kistler JP, et al. The Harvard cooperative stroke registry: a prospective registry. Neurology. 1978;28(8):754.

7. Bogousslavsky J, Van Melle G, Regli F. The Lausanne Stroke Registry: analysis of 1,000 consecutive patients with first stroke. Stroke. 1988;19(9):1083–92.

8. Yiin GSC, Howard DPJ, Paul NLM, Li L, Luengo-Fernandez R, Bull LM, et al. Age-specific incidence, outcome, cost, and projected future burden of atrial fibrillation-related embolic vascular events: a population-based study. Circulation. 2014 Oct;130(15):1236–44.

9. Yurekli UF, Tunc Z. Correlation between Vitamin D, homocysteine and brain-derived neurotrophic factor levels in patients with ischemic stroke. Eur Rev Med Pharmacol Sci. 2022;26(21).

10. Zhou Z, Zhang N, Lin T, Song Y, Liu L, Wang Z, et al. Plasma 25-hydroxyvitamin D3 concentrations and incident risk of ischemic stroke in rural Chinese adults: New insight on ceiling effect. Nutrition. 2022;99:111627.

11. Ohsawa M, Koyama T, Yamamoto K, Hirosawa S, Kamei S, Kamiyama R. 1α, 25-dihydroxyvitamin D3 and its potent synthetic analogs downregulate tissue factor and upregulate thrombomodulin expression in monocytic cells, counteracting the effects of tumor necrosis factor and oxidized LDL. Circulation. 2000;102(23):2867–72.

12. Zhou R, Wang M, Huang H, Li W, Hu Y, Wu T. Lower vitamin D status is associated with an increased risk of ischemic stroke: a systematic review and meta-analysis. Nutrients. 2018;10(3):277.

13. Chan Y-H, Lau K-K, Yiu K-H, Li S-W, Tam S, Lam T-H, et al. Vascular protective effects of statin-related increase in serum 25-hydroxyvitamin D among high-risk cardiac patients. J Cardiovasc Med. 2015;16(1):51–8.

14. Mithal A, Wahl DA, Bonjour J-P, Burckhardt P, Dawson-Hughes B, Eisman JA, et al. Global vitamin D status and determinants of hypovitaminosis D. Osteoporos Int. 2009;20:1807–20.

15. Bouillon R, Norman AW, Lips P. Vitamin D deficiency. N Engl J Med. 2007;357(19):1980–1.

16. Forrest KYZ, Stuhldreher WL. Prevalence and correlates of vitamin D deficiency in US adults. Nutr Res. 2011;31(1):48–54.

17. Yang Q, Churilov L, Fan D, Davis S, Yan B. 1.4 times increase in atrial fibrillation-related ischemic stroke and TIA over 12 years in a stroke center. J Neurol Sci. 2017;379:1–6.

18. Davey Smith G, Ebrahim S. ‘Mendelian randomization’: can genetic epidemiology contribute to understanding environmental determinants of disease? Int J Epidemiol. 2003;32(1):1–22.

19. Revez JA, Lin T, Qiao Z, Xue A, Holtz Y, Zhu Z, et al. Genome-wide association study identifies 143 loci associated with 25 hydroxyvitamin D concentration. Nat Commun. 2020;11(1):1–12.

20. Mishra A, Malik R, Hachiya T, Jürgenson T, Namba S, Posner DC, et al. Stroke genetics informs drug discovery and risk prediction across ancestries. Nature. 2022;611(7934):115–23.

21. Staley JR, Blackshaw J, Kamat MA, Ellis S, Surendran P, Sun BB, et al. PhenoScanner: a database of human genotype–phenotype associations. Bioinformatics. 2016;32(20):3207–9.

22. Kamat MA, Blackshaw JA, Young R, Surendran P, Burgess S, Danesh J, et al. PhenoScanner V2: an expanded tool for searching human genotype–phenotype associations. Bioinformatics. 2019;35(22):4851–3.

23. Hemani G, Zheng J, Elsworth B, Wade KH, Haberland V, Baird D, et al. The MR-Base platform supports systematic causal inference across the human phenome. Elife. 2018;7:e34408.

24. Cui Z, Tian Y. Using genetic variants to evaluate the causal effect of serum vitamin D concentration on COVID-19 susceptibility, severity and hospitalization traits: a Mendelian randomization study. J Transl Med. 2021;19(1):1–13.

25. Nikolakopoulou A, Mavridis D, Salanti G. How to interpret meta-analysis models: fixed effect and random effects meta-analyses. Evid Based Ment Health. 2014;17(2):64.

26. Bowden J, Del Greco M F, Minelli C, Zhao Q, Lawlor DA, Sheehan NA, et al. Improving the accuracy of two-sample summary-data Mendelian randomization: moving beyond the NOME assumption. Int J Epidemiol. 2019;48(3):728–42.

27. Chen J, Yuan S, Fu T, Ruan X, Qiao J, Wang X, et al. Gastrointestinal Consequences of Type 2 Diabetes Mellitus and Impaired Glycemic Homeostasis: A Mendelian Randomization Study. Diabetes Care. 2023;46(4):828–35.

28. Burgess S, Thompson SG. Interpreting findings from Mendelian randomization using the MR-Egger method. Eur J Epidemiol. 2017;32(5):377–89.

29. Zhu X. Mendelian randomization and pleiotropy analysis. Quant Biol. 2020;1–11.

30. Boehm FJ, Zhou X. Statistical methods for Mendelian randomization in genome-wide association studies: A review. Comput Struct Biotechnol J. 2022;20:2338–51.

31. Bowden J, Del Greco M F, Minelli C, Davey Smith G, Sheehan N, Thompson J. A framework for the investigation of pleiotropy in two□sample summary data Mendelian randomization. Stat Med. 2017;36(11):1783–802.

32. Burgess S, Foley CN, Allara E, Staley JR, Howson JMM. A robust and efficient method for Mendelian randomization with hundreds of genetic variants. Nat Commun. 2020;11(1):1–11.

33. Slob EAW, Burgess S. A comparison of robust Mendelian randomization methods using summary data. Genet Epidemiol. 2020;44(4):313–29.

34. Qi G, Chatterjee N. Mendelian randomization analysis using mixture models for robust and efficient estimation of causal effects. Nat Commun. 2019;10(1):1941.

35. Hemani G, Bowden J, Davey Smith G. Evaluating the potential role of pleiotropy in Mendelian randomization studies. Hum Mol Genet. 2018;27(R2):R195–208.

36. Verbanck M, Chen C-Y, Neale B, Do R. Widespread pleiotropy confounds causal relationships between complex traits and diseases inferred from Mendelian randomization. bioRxiv. 2017;157552.

37. Verbanck M, Chen C-Y, Neale B, Do R. Detection of widespread horizontal pleiotropy in causal relationships inferred from Mendelian randomization between complex traits and diseases. Nat Genet. 2018;50(5):693–8.

38. Burgess S, Smith GD, Davies NM, Dudbridge F, Gill D, Glymour MM, et al. Guidelines for performing Mendelian randomization investigations. Wellcome Open Res. 2019;4.

39. Bowden J, Davey Smith G, Burgess S. Mendelian randomization with invalid instruments: effect estimation and bias detection through Egger regression. Int J Epidemiol. 2015;44(2):512–25.

40. Ji W, Zhou H, Wang S, Cheng L, Fang Y. Low serum levels of 25-hydroxyvitamin D are associated with stroke recurrence and poor functional outcomes in patients with ischemic stroke. J Nutr Health Aging. 2017;21:892–6.

41. Huang H, Zheng T, Wang S, Wei L, Wang Q, Sun Z. Serum 25-hydroxyvitamin D predicts early recurrent stroke in ischemic stroke patients. Nutr Metab Cardiovasc Dis. 2016;26(10):908–14.

42. Chaudhuri JR, Mridula KR, Alladi S, Anamika A, Umamahesh M, Balaraju B, et al. Serum 25-hydroxyvitamin d deficiency in ischemic stroke and subtypes in Indian patients. J stroke. 2014 Jan;16(1):44–50.

43. Larsson SC, Traylor M, Mishra A, Howson JMM, Michaëlsson K, Markus HS. Serum 25-Hydroxyvitamin D Concentrations and Ischemic Stroke and Its Subtypes. Stroke. 2018 Oct;49(10):2508–11.

44. Chan Y-H, Schooling CM, Zhao J, Au Yeung S-L, Hai JJ, Thomas GN, et al. Mendelian randomization focused analysis of vitamin D on the secondary prevention of ischemic stroke. Stroke. 2021;52(12):3926–37.

45. Afzal S, Nordestgaard BG. Vitamin D, hypertension, and ischemic stroke in 116 655 individuals from the general population: a genetic study. Hypertension. 2017;70(3):499–507.

46. Lee K-J, Kim H, Lee SJ, Duperron M-G, Debette S, Bae H-J, et al. Causal Effect of the 25-Hydroxyvitamin D Concentration on Cerebral Small Vessel Disease: A Mendelian Randomization Study. Stroke. 2023 Sep;54(9):2338–46.

47. Norman PE, Powell JT. Vitamin D and cardiovascular disease. Circ Res. 2014;114(2):379–93.

48. Tajalli-Nezhad S, Mohammadi S, Atlasi MA, Kheiran M, Moghadam SE, Naderian H, et al. Calcitriol modulate post-ischemic TLR signaling pathway in ischemic stroke patients. J Neuroimmunol. 2023;375:578013.

49. Al Mheid I, Quyyumi AA. Vitamin D and cardiovascular disease: controversy unresolved. J Am Coll Cardiol. 2017;70(1):89–100.

50. Lawlor DA, Harbord RM, Sterne JAC, Timpson N, Davey Smith G. Mendelian randomization: using genes as instruments for making causal inferences in epidemiology. Stat Med. 2008;27(8):1133–63.

